# Target Product Profiles for a Micronutrient Assessment Tool and Associated Blood Collection Device for Use in Population-Based Surveys: An Expert Consensus

**DOI:** 10.1101/2021.05.13.21257124

**Authors:** Emily R. Smith, Joanne Lee, Lindsay H. Allen, David S. Boyle, Eleanor Brindle, Neal E. Craft, Nita Dalmiya, Juergen Erhardt, Dean Garrett, Maria Elena Jefferds, Festo Kavishe, David W. Killilea, Jaqueline K. Kung’u, Anura Kurpad, Cornelia U. Loechl, Sophie E. Moore, Sorrel ML Namaste, Christine M. Pfeiffer, Fabian Rohner, Kerry Schulze, Nazma Shaheen, Sajid Soofi, Pattanee Winichagoon, Bethanie Thomas, Saskia Osendarp, Rahul Rawat

## Abstract

Micronutrient deficiencies are a significant public health problem affecting a large portion of the world’s population. Disproportionately affected populations—infants, young children, adolescents and women of reproductive age including pregnant women — are especially susceptible to the health consequences of insufficient micronutrient intakes. However, assessment of micronutrient deficiencies is not routinely included in population health surveys. This nutrition data gap hampers policy, program, and promotion efforts to prevent and treat micronutrient deficiencies. To address one of the barriers to micronutrient assessment, an expert group created a consensus of a target product profile (TPP) for a micronutrient assessment tool and associated blood collection device for use in population surveys. Experts in laboratory medicine, micronutrient assessment, population-based surveys, and product development reviewed proposed TPP standards and collaboratively established minimum and optimal characteristics. These experts defined the target population as infants and children from 6-59 months, adolescents and women of reproductive age 12-49 years including pregnant women. At minimum, the assessment tool should be a multiplex ELISA formatted for >1 analyte that uses a serum or plasma sample prepared from venous blood obtained by a phlebotomist with a 2-week training. Given the use case was specific for population surveys, experts agreed the minimum tool could be semi-quantitative, with analytical specificity of 99%. The TPP also considers the variable field environments for testing (e.g. storage conditions and time to results). The consensus TPP developed can be used to guide selection of existing technologies into population-based surveys, as well as future investment in product development. Partnerships focused on research and development—including industry, public sector, nonprofit, and academic institutions—can help advance the field and fill the micronutrient data gap.

## Introduction

Globally, micronutrient deficiencies are a major public health problem, and women and children are disproportionately affected by deficiencies and related health consequences (1). Despite effective and low cost interventions to optimize nutritional status including micronutrient supplementation (2) and large-scale food fortification (3), global commitment, funding, and progress toward scaling up these nutrition interventions has been inadequate. Lack of simple, but valid tools for monitoring micronutrient status, is one of the impediments.

Periodic population-based surveys, such as the UNICEF Multiple Indicator Cluster Survey (MICS) and the USAID Demographic and Health Surveys (DHS), remain the cornerstone of national and sub-national nutrition data systems. However, micronutrient assessment in these surveys remains limited, and very few countries have implemented specific micronutrient surveys. The MICS does not currently include any micronutrient biomarker analysis. Sometimes micronutrient status is estimated based on dietary self-reports or limited to a single biomarker. For example, many surveys routinely collect information on anemia (based on hemoglobin measurement) and intakes of vitamin A and iron-rich foods. The DHS has previously, on occasion, included micronutrient biomarkers (i.e. transferrin receptor, serum retinol, retinol binding protein, and C-reactive protein) to assess iron and vitamin A deficiency at the population level; these efforts add to the technical, logistical, and financial burden (4). In a few other cases, the DHS has also partnered with local organizations to administer micronutrient assessments, for example in Cambodia and Malawi (5, 6).

Although collection of micronutrient biomarker data is increasingly common, getting representative data on micronutrient status across a range of populations at national and subnational level remains a challenge. These data gaps are barriers to quantifying the problem, developing evidence-based policy, targeting resources, and ensuring accountability (7). Barriers include the costs of specimen collection, the complexity posed by household-based sample collection for biomarker assays, cold chain, sample transportation and storage, the laboratory infrastructure or capacity required to conduct these tests, cost for analysis, and a limited number of commercially available, valid, and reliable assays. The Biomarkers for Nutrition Development (BOND) and Biomarkers Reflecting Inflammation and Nutritional Determinants of Anemia (BRINDA) projects have made major advances in consolidating our current understanding related to measuring and interpreting micronutrient status for a number of key micronutrients (8–14); (15, 16). There have also been some limited recent advances in developing new micronutrient assessment tools (17–21). However, uptake has been limited in population health surveys to date. Many point to lack of validation against gold standard methods, lack of external validation, or mixed results from external validation, as important barrier to uptake. Further, we hypothesize that both limited investment in development of novel micronutrient assessment tools and limited uptake of new tools is partly due to a lack of consensus in the field around the necessary technical specifications of micronutrient assessment tools required to overcome current barriers to broader inclusion in population health surveys.

Here we describe a consensus building exercise focused on the development of a target product profile (TPP) for a micronutrient assessment tool and associated blood collection device. TPPs are often part of a strategic planning process to guide research and development related to drug or diagnostics discovery. A diverse panel of global health, nutrition science, and survey implementation experts created TPPs based on a systematic, iterative process. The development of TPPs for a micronutrient assessment tool and associated blood collection device for use in population health surveys is intended to demonstrate areas of consensus. Ideally, the TPPs will help drive investment in new tools ensuring that end-user and market needs are met. Given the complexity of population surveys and existing barriers around micronutrient biomarker assessment, simple and effective new tools are needed.

## Materials and Methods

### Description of the TPP

A TPP is a tool which has been used widely to support and guide diagnostic development in various fields of health (22). This project was initiated by nutrition experts at the Bill & Melinda Gates Foundation (BMGF) and the Micronutrient Forum, and the developed TPPs are intended to provide guidance for funders, as well as researchers in the public and private sector, for prioritizing investments, research, and development of new micronutrient assessment tools for use in large-scale surveys. The TPPs are broad, and thus not specific to measurement of any particular micronutrient. Nor does the TPP imply that a single tool should be developed to assess the entire breadth of micronutrients that might be measured in a population health survey. The TPPs can also be used to assess the fit of emerging tools for use in population health surveys.

For each field of the TPP, there were “minimal” and “optimal” standards provided. Minimal standards were defined as characteristics which provide the best possible practice given existing technology, resources, and market conditions. They were further clarified in the multiple rounds of input as “must have” characteristics – that is, minimal characteristics that are feasible under current conditions and constitute the baseline threshold which micronutrient assessments must meet when being included in population health surveys. Optimal standards were defined as “nice to have” and consisted of desired attributes which are best suited to assess individual, population health, and survey administrator needs which may not yet be achieved given the limitations of existing technology and market conditions.

To better reflect prioritization of micronutrients to be included in the tool, we labeled micronutrients as high- and low-priority for the optimal TPP. Experts agreed that these additional categories provided a meaningful distinction in ranking relative importance of included micronutrients, although this departs from the traditional structure of a TPP.

### Modified Delphi Approach

We developed the TPPs using a modified Delphi approach (23, 24). The Delphi method provides a systematic way of eliciting and summarizing expert opinion. Briefly, the lead investigators (ES, RR, SO, JL, BT) drafted initial TPPs based on professional experience and a literature review conducted in April 2019. The expert panel (LHA, DSB, EB, NC, ND, OD, JE, DG, MEJ, FK, DK, JKK, AK, JL, CUL, SEM, SN, CMP, FR, KS, NS, SS, BT, PW) provided feedback and proposed modifications to the TPPs through two rounds of anonymized, online feedback in May 2019. In June 2019 we conducted a third round of in-person discussion and modification in Baltimore, MD, USA (held in conjunction with the annual meeting of the American Society for Nutrition).

The TPP template was based on existing TPPs and assessment parameters developed by experts at PATH and the University of Washington (25, 26), as well as general templates developed across a range of product development experience provided by the Bill & Melinda Gates Foundation. Initial values were included in the TPP based upon published values of commonly used micronutrient assays (20), a recently developed micronutrient assay (18), and other published TPPs for diagnostics for use in low and middle-income countries (LMICs) (23). In cases where there was no available information, the TPP field was left blank.

The expert working group reviewed the first draft TPPs individually via an online survey (S1 File, S2 File). For each TPP characteristic, experts rated their agreement with the proposed standards (1-strongly disagree to 5-strongly agree, or “Decline to Answer”) and provided written justification for their opinion. Fields which achieved the *a priori* stop criteria of ≥75% agreement (having responded “agree” or “strongly agree”) were considered having achieved expert consensus and closed for further revision and comment in subsequent rounds of TPP review. Fields that did not achieve consensus during Round One were revised based on expert comments, and a second draft of both TPPs were generated.

The second draft of both TPPs were distributed individually via an online survey (S3 File, S4 file), including anonymized comments from the first round to accompany revisions in fields which did not achieve consensus in the first round of review. These anonymized comments were included to help inform working group participants, who as previously described included a wide range of expertise. Experts were encouraged to revise their earlier recommendations based on perspectives shared in the first round of feedback, with the intent to drive agreement to ≥75% on minimal and optimal standards. Fields that did not obtain consensus during Round Two were revised based on expert comments, and a Round Three drafts of the TPPs were generated, again including anonymized comments (S5 file, S6 file).

Fields that did not achieve consensus during Round Two were prioritized for discussion during the in-person convening based on discordance of opinion. Proposed revisions and compiled comments were shared with the expert panel in advance of the in-person convening and then discussed, with a third round of review, evaluation, and discussion on the latest TPP drafts. Discussion points and third-round live voting on outstanding TPP standards for alignment were documented, and final TPPs were established after achieving ≥75% agreement by the in-person group.

### Expert Group

The expert working group consisted of individuals from academic institutions, population health survey-implementing organizations, governments, international organizations, product development partners, industry partners, and for-profit organizations. Twenty-three people were invited to join the expert working group and 22 agreed to participate. The TPP expert working group members were nominated by ERS, RR, and SO based on nominees’ expertise as recognized leaders in the field of micronutrient assessment, laboratory science, population health surveys, global health, and product development. To gain representative and balanced feedback from the working group, experts from a range of geographies including North America, Europe, Asia, and Africa and professional backgrounds were selected. Expert working group members were not aware of the identities of other members of the working group until the June 2019 convening.

## Results

The surveys were split into two parts: “Blood Collection Device” and “Micronutrient Assessment Tool.” During Round One review (n=21 participants), the group achieved clarity and expert consensus on 12 of 29 fields in the micronutrient assessment tool TPP. In Round Two (n=19 participants), one new field was added to the TPP based on feedback, and we achieved consensus on 12 of the remaining 18 fields. We achieved consensus on the remaining six fields during the third round of review with 18 experts joining at the June 2019 convening (Fig 1). Additional minor, clarifying edits were made during the manuscript preparation process.

**Figure 1.**
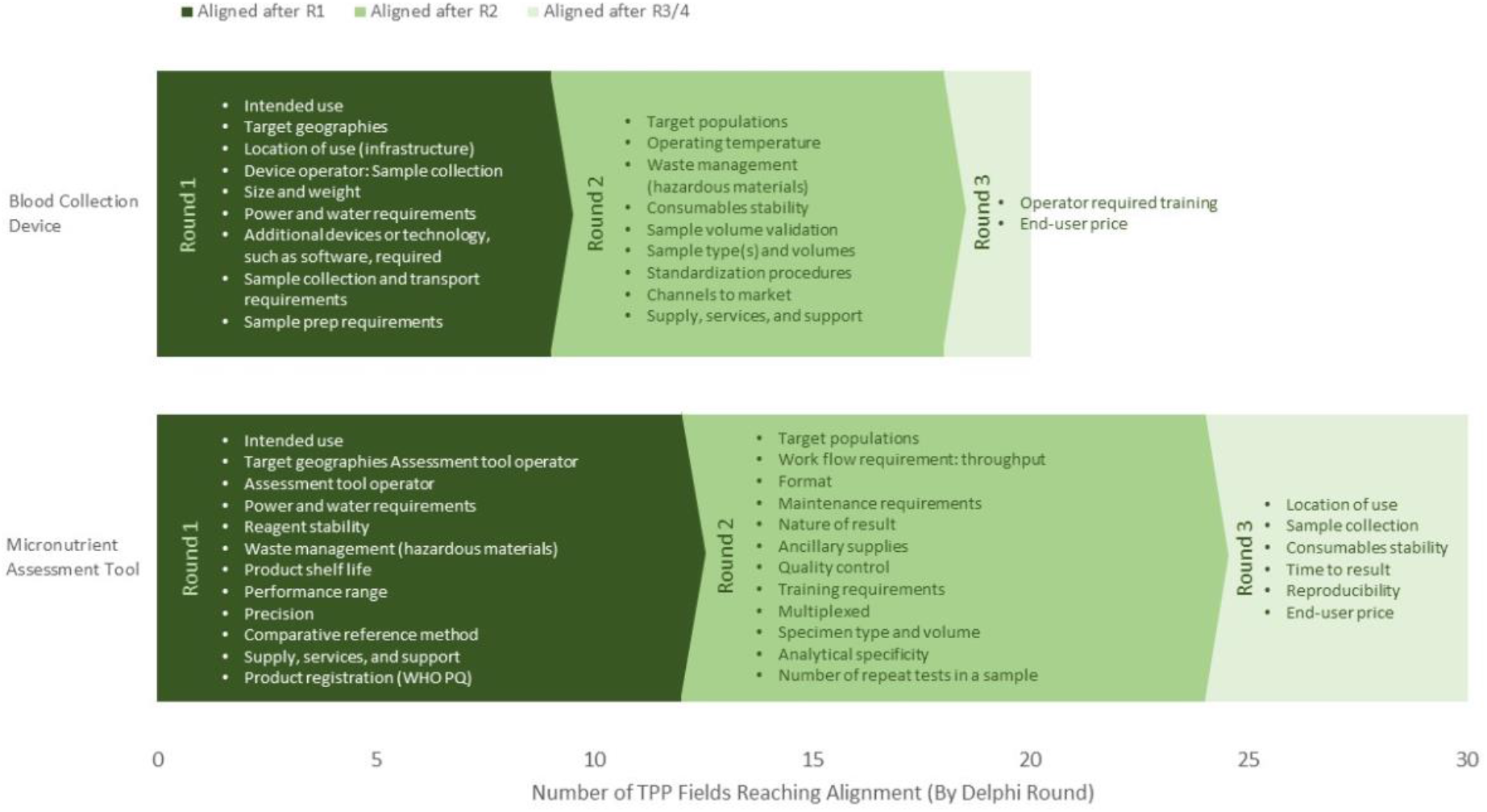
Micronutrient Assessment Tool Target Product Profile (TPP) and Blood Collection Device TPP Fields Reaching Alignment, Stratified by Delphi Round.

Regarding the TPP for blood collection device, in Round 1 we achieved clarity and expert consensus on 9 of 23 fields. Two fields (“sterilization requirements” and “types of specimens/assays run in the same facility”) were eliminated from the TPP based on expert feedback. During Round 2, we achieved consensus on 9 of the remaining 12 fields, and one field was eliminated (“Work flow requirement: throughput”). The final two fields were discussed during the June 2019 convening (Fig 1).

During the June 2019 convening (i.e. Round Three), we discussed the following high priority fields: sample collection (micronutrient assessment tool), end-user price (micronutrient assessment tool and blood collection device), and specific micronutrient categorization. High priority fields were determined by level of discordance as well as discordance of recommendations and comments in the previous two rounds of review.

Remaining fields without expert consensus were deemed low priority for discussion because there was a clear path to alignment based on Round Two comments (e.g. operator required training, location of use, consumable stability, time to result, reproducibility). These revised standards were shared with the full expert working group in advance of the convening for their review and comment as well; however, structured time for polling evaluation of these fields was not provided given time constraints.

The final micronutrient assessment tool TPP is provided in Table 1, and the final blood collection device TPP is provided in Table 2.

**Table 1.**
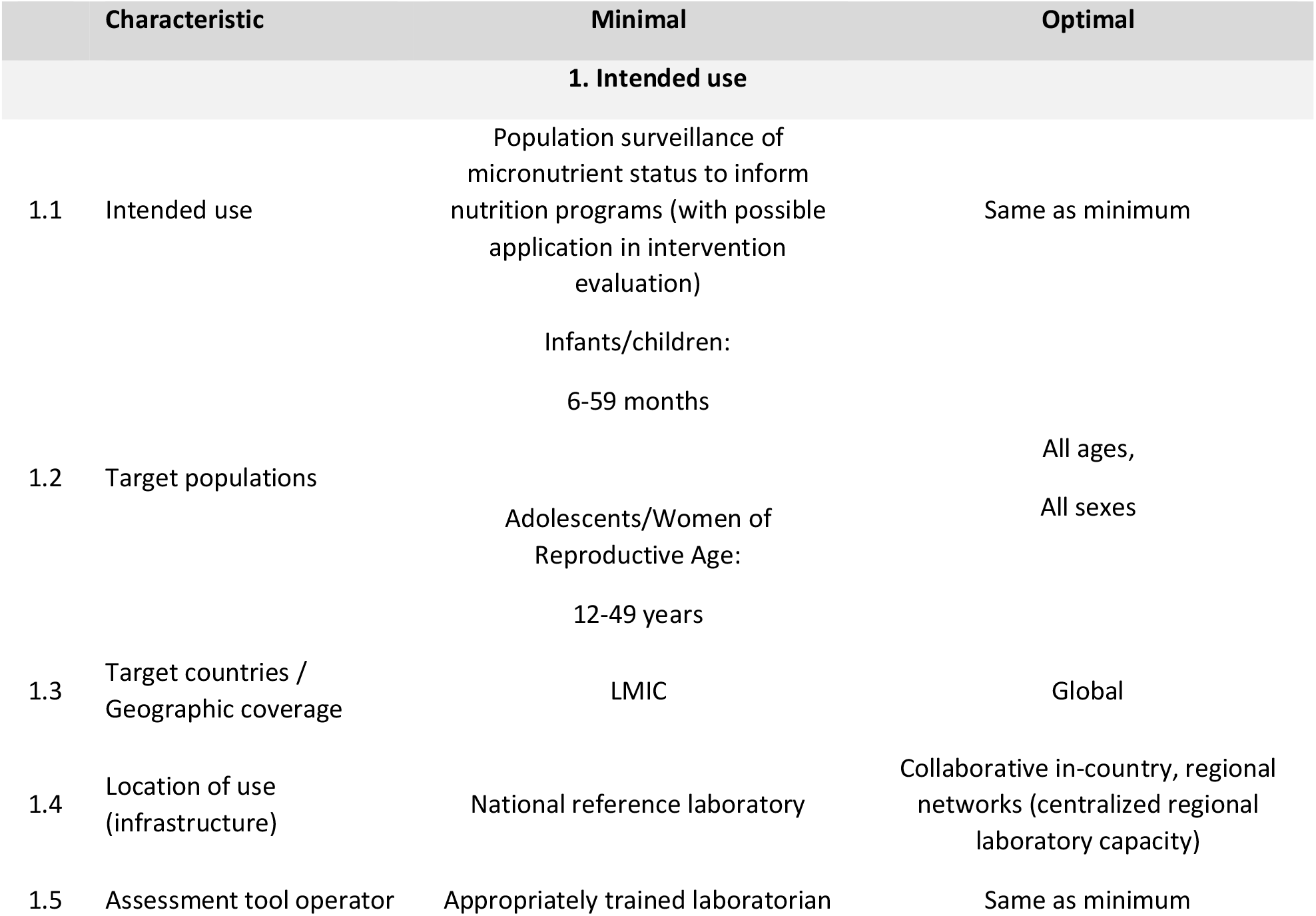
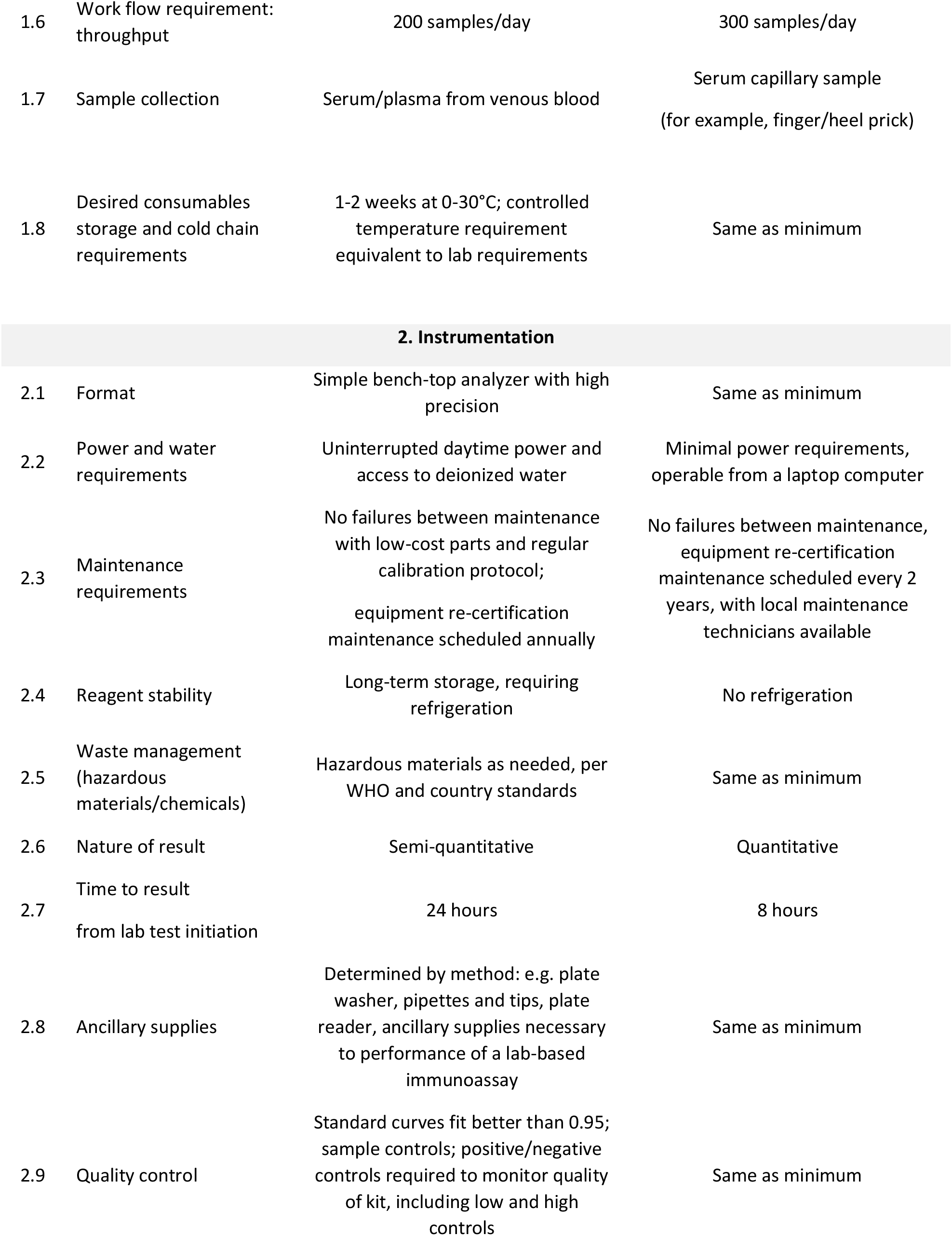
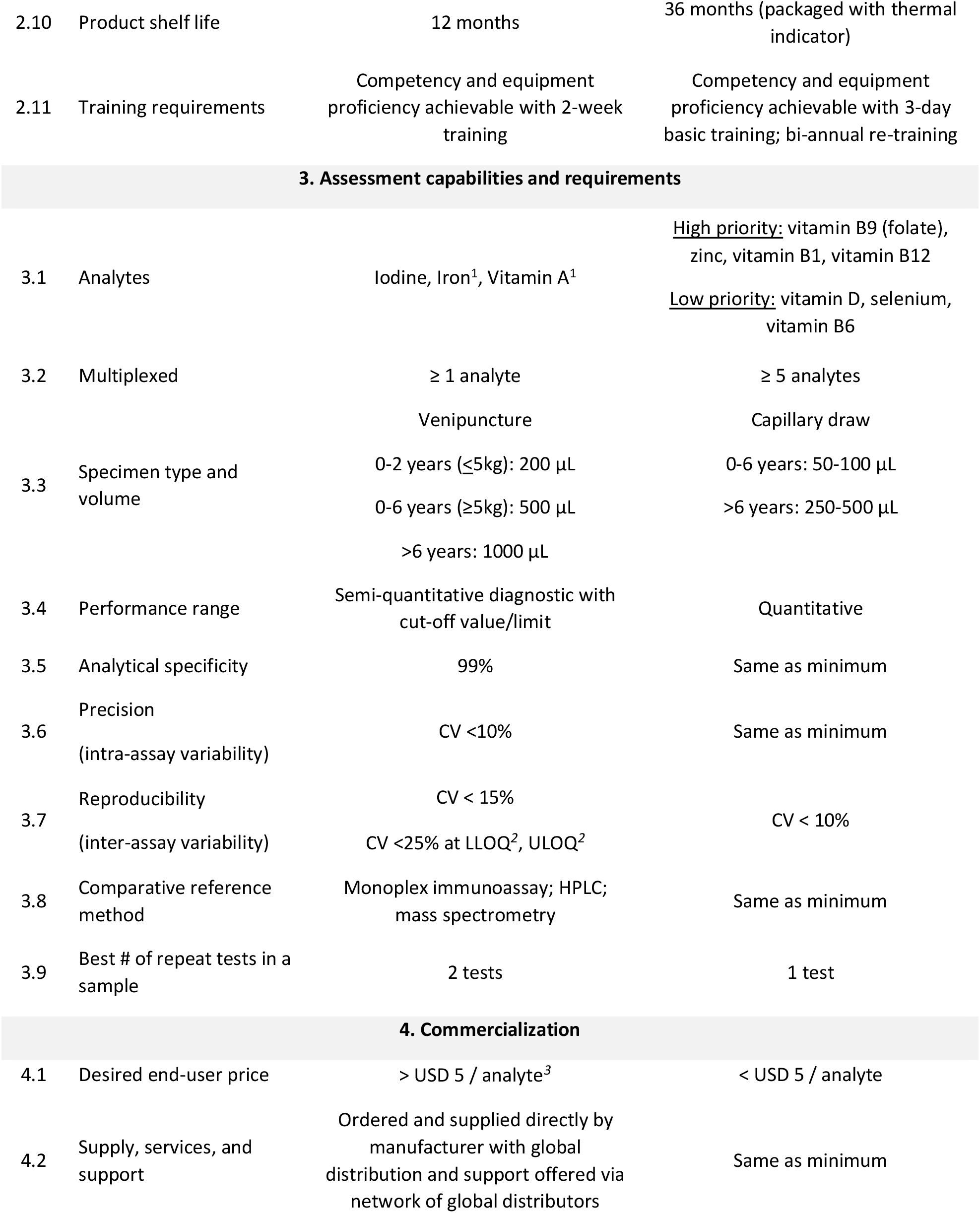
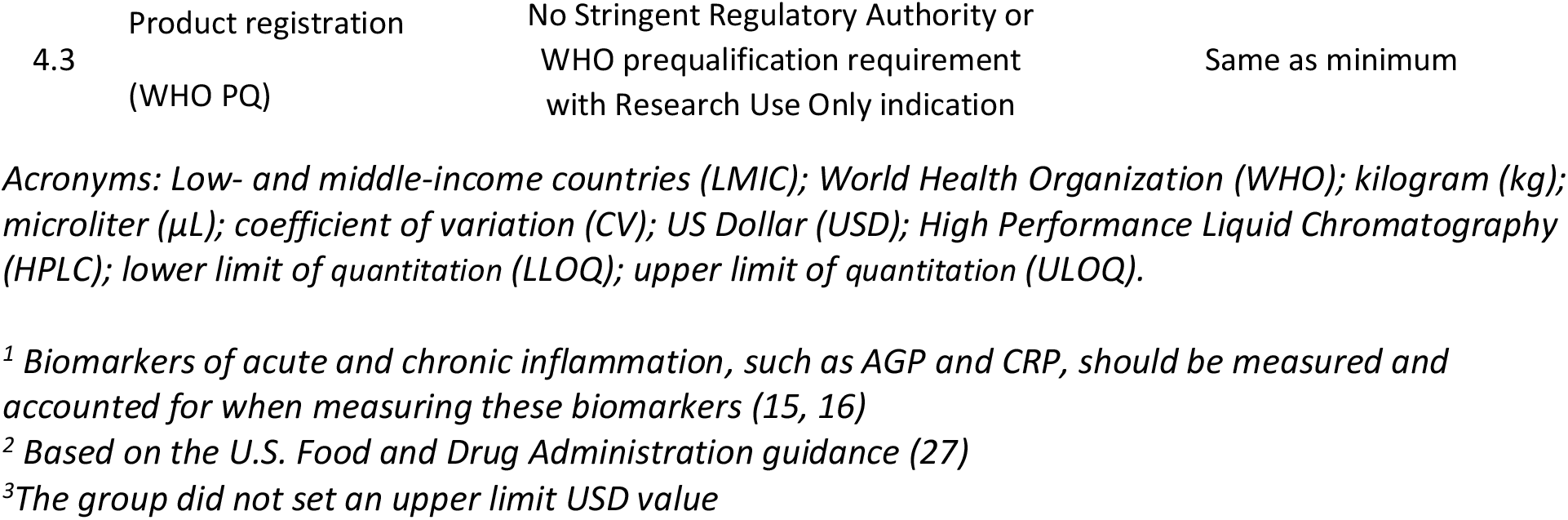
Micronutrient Assessment Tool for Population-Based Surveys Target Product Profile: Minimal and optimal characteristics.

**Table 2:** Blood Collection Device for Population-Based Surveys Target Product Profile (TPP): Minimal and optimal characteristics.

|  | Characteristic | Minimal | Optimal |
| --- | --- | --- | --- |
| **1. Intended use** |  |  |  |
| 1.1 | Intended use | Population surveillance of micronutrient status to inform nutrition programs (with possible application in intervention evaluation) | Same as minimum |
| 1.2 | Target populations | Infants/children: 6-59 months<br>Adolescents/ Women of Reproductive Age: 12-49 years | All ages,<br>All sexes |
| 1.3 | Target countries / Geographic coverage | LMIC | Global |
| 1.4 | Location of use (infrastructure) | Household / field use | Same as minimum |
| 1.5 | Device operator: Sample collection | Trained phlebotomist | Trained household survey worker |
| **2. Device characteristics** |  |  |  |
| 2.1 | Size and weight | Handheld | Same as minimum |
| 2.2 | Power and water requirements | None | Same as minimum |
| 2.3 | Operating temperature | 15-35°C, 35-85% humidity | Same as minimum |
| 2.4 | Waste management (hazardous materials/chemicals) | No hazardous materials or chemicals required for blood collection device; same as standard phlebotomy equipment | No hazardous materials or chemicals required for blood collection device<br>Single use and safe disposal mechanism, same as standard phlebotomy equipment |
| 2.5 | Consumables stability | Stable at ambient temperature and humidity for 3 months | Stable at ambient temperature and humidity for 6 months |
| 2.6 | Additional devices or technology, such as software, required | None required | Same as minimum |
| 2.7 | Sample volume validation | Clear indicator to confirm sufficient volume collected | Same as minimum |
| **3. Sample handling** |  |  |  |
| 3.1 | Sample type(s) and volumes | Venous blood draw<br>0-2 years (< 5kg): 200 µL<br>0-6 years (≥ 5kg): 500 µL<br>>6 years: 1000 µL | Capillary blood draw<br>0-6 years: 50-100 µL<br>>6 years: 250-500 µL |
| 3.2 | Sample collection and transport requirements | Cold chain required (0-4° C) | Ship without cold chain; should tolerate stress during transport |
| 3.3 | Sample prep requirements | In-field centrifugation and aliquot required | In-field processing not required |
| 3.4 | Operator parameters, required training | Operator training limited, e.g. 1 day of training by competent field staff | Same as minimum |
| 3.5 | Standardization procedures | Lab facility has internal standardization | In-country, regional or global facility is accredited to conduct standardization |
| **4. Commercialization** |  |  |  |
| 4.1 | Desired end-user price of device sampling | > \$ 5 USD / sample<sup>1</sup> | < \$2 USD / sample |
| 4.2 | Channels to market | Mainstream regional laboratory suppliers | Mainstream regional and local laboratory suppliers |
| 4.3 | Supply, services, and support | Does not require special services nor support | Same as minimum |

Acronyms: Low- and middle-income countries (LMIC); World Health Organization (WHO); kilogram (Kg); microliter (µL); US Dollar (USD).
<sup>1</sup>The group did not set an upper limit USD value

## Discussion

The TPPs for the micronutrient assessment tool and the associated blood collection device reflect the consensus of a group of key stakeholders and experts and are intended to guide future development of relevant tools. Clearly defined standards established through a systematic agreement process of a diverse group of global experts can help advance technological innovation. The TPPs can also aid in decision-making to adopt new tools and to invest in product development.

### Micronutrient assessment tool end-user costs

Inclusion of micronutrient assessment in population health surveys requires available and scalable tools; assay costs currently present one of the significant barriers to achieving this. While the lowest cost biomarker analyses for a few analytes are available at an approximate cost of $1 USD/analyte (20), the current contract laboratory infrastructure does not support more than a few micronutrient surveys in a given year and is unlikely to be able to support further growth in the market. Furthermore, multiple experts report that despite significant internal investment, they have been unable to develop similar assays at this low cost given the limited selection and high costs of commercially available antibodies. The recently developed Q-Plex™ is available at $1.60/analyte for LMICs, but has not yet been widely used in the population survey context, and is currently limited to only a few biomarkers of micronutrient status. Panel experts report that other existing assay costs range widely depending on institutional differences in personnel costs and overheads, micronutrient and biomarker analytes in question, and analytical technology used. For example, ferritin (as a biomarker for iron status) is often quoted in the $10-45 USD range, serum retinol-binding protein (RBP, a vitamin A biomarker) in the $15-40 USD range, and other biomarkers such as serum retinol by HPLC in the $25-60 USD range.

There was a wide range of opinions about the minimal and optimal costs of these products. Experts noted that it would be important to understand the broader market dynamics to reasonably define a minimum and optimal end-user price for micronutrient assessment. We set the optimal standard at <$5 USD/analyte because it was the lowest cost where there was some agreement among implementers and laboratory product development experts that this price point both seemed “low cost” and that it would be potentially feasible to make such a product. Open-ended cost standards were established in consideration of commercially available assessment tools. However, panelists agreed that further research is needed to understand consumer willingness to pay, total projected demand volume (or opportunities to create demand through advocacy work), and to obtain a deeper understanding of associated assessment costs (e.g. training, infrastructure and labor savings with new technological innovation, and quality of available technologies).

### Considering point-of-care diagnostics

When considering the optimal standards for a micronutrient assessment tool for population health surveys, experts discussed whether this should be a laboratory-based or point-of-care assessment tool that could be used in the household by trained survey administrators. Several experts argued in favor of a point-of-care tool because it would eliminate the complexity of transporting or shipping biological samples to labs, and could improve timeliness in disseminating and using relevant data to assist in policy and program decisions. Further, they noted the potential benefits of sharing micronutrient status results while still in the field, especially when the data has clinical relevance (e.g. iron deficiency anemia). However, most experts pointed out that it is not realistic to achieve a point-of-care tool which fulfills the complete list of other requirements of the micronutrient assessment tool TPP (e.g. semi-quantitative, multiplexed), given the current technological barriers and cost. Point-of-care technology may add complexity in equipment transport, harmonization, household setup, quality control, external quality assurance, and results interpretation. In fact, some experts argued that household-based assessment might create undue burden on survey administrators or even the health system; protocols and a standard of care regarding who would be referred for treatment, when, and for what treatment would need to be developed in collaboration with local governments. Furthermore, a laboratory-based tool was deemed preferable for reasons such as the efficiency of lab-based methods and also the additional depth of information which can be provided through laboratory-based tests (e.g. quantitative results). Given the intended use in large-scale population health surveys and technological limitations, the TPP outlined assumes an optimal preference for a reliable and user-friendly laboratory-based micronutrient assessment tool. While clinical application was not the focus of this activity, a point-of-care assessment tool is still a critical gap in low resource clinical settings, especially for assessment of iron status.

### Other barriers to micronutrient assessment

The expert working group identified several other barriers to measuring micronutrient status in populations-based surveys, including a lack of comparative reference methods, standardized reference analytes (e.g. soluble transferrin receptor (sTFR) and RBP), and lack of high-quality, commercially available antibodies for some micronutrient biomarkers (e.g. sTfR or holotranscobalamin). An effort to prioritize available biomarkers for each micronutrient was not part of this process, and future efforts to do so may help guide those wishing to include micronutrient assessment in population health surveys. Development of this more detailed guidance will require more in-depth discussions by technical expert working groups familiar with the available scientific data on existing biomarkers for population-based assessment. Previously, the Inflammation and Nutritional Science for Programs/Policies and Interpretation of Research Evidence (INSPIRE) and BRINDA working groups highlighted the importance of considering inflammation in the measurements and interpretation of micronutrient assessment (15, 16). In addition, experts pointed to limited existing reference laboratories globally as a key barrier. Some experts suggested that a market analysis—providing a deeper understanding of the potential size of the market for this type of technology—would allow for insight into the best approach toward solving these barriers. Furthermore, clear understanding of the market can accelerate academic and private sector investment in research and development of new tools.

### Strengths and limitations

A key strength of the final TPPs are that they were generated through the systematic Delphi approach– established *a priori*– to create a consensus-driven final product. Our expert panel included a diverse range of expert stakeholders, intentionally including variety in content expertise, institutions/sectors, and geographies. Additionally, the survey and convening process provided an opportunity for iteration and information sharing both anonymously and through discussion. However, the consensus TPPs included here are the reflection of the opinion of this single group of expert stakeholders and may not reflect all expert opinions. Further, due to scheduling and logistical barriers (e.g. time zone differences), the June 2019 convening included only a subset of the full group, and thus not all experts participated in the final round, which was the only round with opportunity for discussion and debate among experts. However, all expert panel members have reviewed the final TPPs and this manuscript.

While micronutrients were categorized as minimal and optimal in the TPPs, we did not include details about micronutrient-specific biomarkers for micronutrient assessment in these TPPs for several reasons. First, there has been significant expert work regarding biomarkers through the BOND and New York Academy of Science reviews of specific micronutrients (8–14, 28, 29); these reports provide a thorough review of the science related to potential biomarkers for each micronutrient. Second, assay limitations may restrict which biomarkers can be multiplexed together. Thus, new tools may require tradeoffs between biomarker choice for a given micronutrient in order to create a multiplex tool.

## Conclusion

The TPPs presented in this paper demonstrate broad agreement among select key stakeholders for both micronutrient assessment tools and associated blood collection devices for specific use in population health surveys. The current document can help guide development of innovative assessment platforms that identify micronutrient deficiencies at a population level and thereby guide resource allocation and program design to effectively address micronutrient deficiencies globally. There could be wide application of new tools in population-based health surveys and other research or surveillance efforts, and as such, the potential impact of new tools is large. The expert working group agreed that development of these generic TPPs are a first step towards overcoming some of the current perceived technical and logistical barriers to improving the availability of high-quality micronutrient status data from population surveys. The publication of these TPPs is intended to facilitate the development and application of measurement tools and lead to efforts to prioritize biomarkers, develop comparative reference standards, and quantify the market value of future innovations. The gap in micronutrient status data is well documented and thus, the burden of micronutrient deficiencies among women and children has gone largely unaddressed and is under-resourced. These TPPs establish a benchmark for new tools, which can guide product development work. We consider this to be one critical step in filling the nutrition data gap.

## Data Availability

All data is included in the manuscript and supplementary files.

## Acknowledgments

The authors would like to acknowledge and thank Shauna Hargrove (Bill & Melinda Gates Foundation) for her administrative support through the process, and both Shauna Hargrove and Rebecca Schreiner (Opus Agency) for event planning and coordination support around the June 2019 convening. We acknowledge Dr. Sabine Dittrich (Foundation for Innovative New Diagnostics) as well for sharing her previous experience with the application of a modified Delphi process for TPP development. We acknowledge Omar Dary (United States Agency for International Development) for sharing his expertise in improving this work. We also thank Siran He (George Washington University) for her editorial assistance.

## Author contributions

**Conceptualization:** JL, SO, RR, ES, BT

**Formal analysis:** JL, ES, BT

**Funding Acquisition:** RR, ES

**Participated in Delphi Consensus Process:** LHA, DSB, EB, NC, ND, OD, JE, DG, MEJ, FK, DK, JKK, AK, JL, CUL, SEM, SN, CMP, FR, KS, NS, SS, BT, PW

**Methodology:** ES, RR, SO, BT, JL

**Project Administration:** ES, BT, JL

**Supervision:** ES, RR, SO, BT, JL

**Writing – Original Draft Preparation:** ES, RR, SO, JL

**Writing – Review & Editing:** LHA, DSB, EB, NC, ND, OD, JE, DG, MEJ, FK, DK, JKK, AK, JL, CUL, SEM, SMLN, CMP, FR, KS, NS, SS, BT, PW

