## Supplementary File 1 for "Target Product Profiles for a Micronutrient Assessment Tool and Associated Blood Collection Device for Use in Population-Based Surveys: An Expert Consensus"

**Supplementary File 1 (S1). Round 1 Micronutrient Assessment Tool Target Product Profile (TPP) draft reviewed by the expert panel, spring 2019**

### I. Background

While micronutrient (MN) malnutrition is highly associated with adverse health outcomes and there have been several global investments to develop tools^[1]^ to assess distribution of MN deficiencies and understand the magnitude of the problem, uptake of such assessment tools by population health surveys has remained low.

MN assessment in population health surveys is complicated by a number of challenges ranging from those related to field-based data collection, laboratory infrastructure, and the absence of quality assurance programs as well as globally standard reference assays. To date, the Demographic and Health Surveys (DHS) have on occasion included transferrin receptor, retinol serum, and C-reactive protein to assess iron and Vitamin A deficiency at the population level, efforts which required investment in local infrastructure (Garrett, et al. 2011). The UNICEF Multi-Indicator Cluster Survey (MICS) does not currently include any micronutrient biomarker analysis.

The lack of consensus in the field around the best practices for micronutrient assessment to meet this use case has been identified as a barrier to wider uptake by population health surveys. The development of a high-level, conceptual TPP for an MN assessment tool for use in population health surveys is intended to address this barrier as a necessary precursor for efforts to broaden use.

### II. Critical assumptions

This target product profile (TPP) is intended to serve as a high level “conceptual” TPP to build field consensus around the minimum and optimistic characteristics required for a micronutrient assessment tool to be used in population health surveys. As such, the TPP is not designed to point specifically to any one existing assessment methodology or product. The predefined TPP characteristic categories below will serve to guide the field in assessing fit of existing micronutrient assessment tools and, in the absence of existing technologies to meet minimum TPP guidelines, investment in the development of appropriate assessment tool.

While we acknowledge there will be supply chain and personnel requirements for this device to be successful (e.g., equipment maintenance and repair, available replacement parts), we will not explicitly address how to resolve these challenges in this TPP.

### III. Target product profile (TPP) overview**^[2]^**

Please refer to **Appendix A** for specific questions referring to content below in addition to overall content review.

|  | **Characteristic** | **Minimum (“must”)^[3]^** | **Optimal (“want”)^[4]^** | **Annotation** |
| --- | --- | --- | --- | --- |
| **1** | **Intended use** | | | |
| 1.1 | Intended use | Population surveillance of micronutrient deficiency to inform nutrition programs | Same as minimum | (PATH 2017) |
| 1.2 | Target populations | Infants/children: 6-59 months  Women of reproductive age (WRA): 15-45 years | All ages | (PATH 2017) |
| 1.3 | Target countries / Geographic coverage | Low-income or middle-income country (LMIC) | Global | (PATH 2017) |
| 1.4 | Location of use (infrastructure) | National reference laboratory | District-level laboratory | (PATH 2017) |
| 1.5 | Assessment tool operator | Laboratorian | Same as minimum | (PATH 2017) |
| 1.6 | Work flow requirement: throughput | 400 samples/day | 1000 samples/day | (PATH 2017) |
| 1.7 | Sample collection | Serum/plasma from venous blood | Finger/heel-prick blood-derived dried blood spot and/or serum/plasma spot | (PATH 2017) |
| 1.8 | Desired consumables sample stability, storage, and cold chain requirements | 6 months at fluctuating temperatures (0-40°C);  <90% relative humidity;  no controlled temp. required | 12 months at fluctuating temperatures (0-40°C);  No controlled temp. required;  no cold chain requirement | (BMGF 2018) |
| **2** | **Instrumentation** | | | |
| 2.1 | Format | Enzyme-linked immunosorbent assay (ELISA) | ELISA |  |
| 2.2 | Power and water requirements | Uninterrupted power and access to deionized water | Battery-powered with minimal power requirements, operable from a laptop computer; plate shaker can use universal power supply | (PATH 2017) |
| 2.3 | Maintenance requirements | No failures between maintenance, with maintenance scheduled annually | No failures between maintenance, with maintenance scheduled every 5 years | (PATH 2017) |
| 2.4 | Reagent stability | Long-term storage, requiring refrigeration | No refrigeration | (PATH 2017) |
| 2.5 | Waste management (hazardous materials/chemicals) | Hazardous materials as needed, per the World Health Organization (WHO) and country standards | Same as minimum | (PATH 2017) |
| 2.6 | Nature of result | Qualitative;  Yes/No (deficient) indicator | Quantitative | (BMGF 2018, PATH 2017) |
| 2.7 | Time to result | 4 hours |  | (PATH 2017) |
| 2.8 | Ancillary supplies | Plate washer, pipettes and tips, other standard ancillary supplies necessary to performance of a lab-based immunoassay | Same as minimum | (PATH 2017) |
| 2.9 | Quality control | Standard curves fit better than 0.98; sample controls; positive/negative controls required to monitor quality of kit | Same as minimum | (PATH 2017, Dittrich, et al. 2016, Erhardt, Estes, et al. 2004) |
| 2.10 | Product shelf life | 12-18 months | 36 months (packaged with thermal indicator) | (PATH 2017) |
| 2.11 | Training requirements | **Recommendation needed** | **Recommendation needed** |  |
| **3** | **Assessment capabilities / requirements** | | | |
| 3.1 | Analytes |  |  |  |
| 3.2 | Multiplexed | ≥ 1 analyte | ≥ 2 analytes | (BMGF 2018) |
| 3.3 | Specimen type and volume | Venupuncture  0-2 years: 70-100 µL  2-6 years: 50-100 µL (~2 drops)  >6 years: 250-500 µL (~4 drops) | Dried blood spot / matrix  10-40 µL (~1 drop) | (BMGF 2018) |
| 3.4 | Analytical limit of detection | Limit of detection should be such that it allows clinically relevant performance as defined below | Same as minimum | (Dittrich, et al. 2016) |
| 3.5 | Analytical specificity | 99% | Same as minimum | (PATH 2017) |
| 3.6.1 | Precision  (intra-assay variability) | 95% SD between repeats | Same as minimum | (OIE 2013) |
| 3.6.2 | Reproducibility  (inter-assay variability) | CV < 15% | CV < 7.5 | (BMGF 2018, Erhardt, Estes, et al. 2004) |
| 3.7 | Comparative reference method | Monoplex immunoassay; HPLC; mass spectrometry | Same as minimum | (PATH 2017) |
| **4** | **Commercialization** | | | |
| 4.1 | Desired end-user price | 5 USD / analyte | 1 USD / analyte | (BMGF 2018, PATH 2017) |
| 4.2 | Supply, services, and support | Ordered and supplied directly by manufacturer with global distribution and support offered via network of global distributors | Same as minimum | (PATH 2017) |
| 4.3 | Product registration (WHO PQ) | No Stringent Regulatory Authority (SRA) or WHO prequalification requirement with Research Use Only indication | Same as minimum | (PATH 2017) |

### [TPP 3.1 REFERENCE] Table 1: Established Biomarkers for MNDs^[5]^

| Nutrient | Biomarker | Coding |
| --- | --- | --- |
| Calcium | Not measured | m: metabolite  e: element  p: protein  f: functional  b: buffered (no signal until severe deficiency)  u: unusual  n: non-specific |
| Choline | Plasma choline (m,u,b) |  |
| Iodine | Urine iodine (m),  Plasma thyroglobulin (p) |  |
| Iron | Plasma ferritin (p)  Plasma soluble transferrin receptor (p)  Plasma hepcidin (p),  Plasma transferrin (p) |  |
| Selenium | Plasma SEPP1 (p,u),  Plasma selenium (e,u) |  |
| Vitamin A | Plasma retinol (m,b),  Plasma RBP4 (p,b) |  |
| Vitamin B1 (thiamine) | Plasma thiamine/TMP (m),  Whole blood thiamine/TMP (m),  RBC ThDP (m),  RBC transkelotase (f) |  |
| Vitamin B3 (niacin) | Plasma niacin/nicotinamide (m) |  |
| Vitamin B6 (PLP) | Plasma PLP/PA (m) |  |
| Vitamin B9 (folate) | RBC folate (m),  Plasma folate (m) |  |
| Vitamin B12 (cobalamin) | Plasma cobalamin (m),  Plasma holotranscobalamin (p),  Plasma homocysteine (m,n),  Plasma methylmalonic acid (m,n) |  |
| Vitamin D | Plasma 25-OH-D |  |
| Vitamin E | Plasma α-tocopherol (m) |  |
| Vitamin K | PIVKA (p,u) |  |
| Zinc | Plasma zinc (e,b) |  |
| Other B vitamins | Plasma metabolites |  |

### References

BMGF. 2016. "Diagnostics Assay/Instrument - Target Product Profile: Serum Bilirubin Monitor and Non-invasive Bilirubin Monitor."

BMGF. 2018. "Generic Target Product Profile for Micronutrient Biomarker for Population-Based Surveys."

Brindle, Eleanor. 2014. "Biomarkers for Vitamins and Minerals." Center for Studies in Demography and Ecology, University of Washington.

Dittrich, S, BT Tadesse, F Moussey, A Chua, A Zorzet, T Tangden, DL Dolinger, et al. 2016. "Target Product Profile for a Diagnostic Assay to Differentiate between Bacterial and Non-Bacterial Infections and Reduce Antimicrobial Overuse in Resource-Limited Settings: An Expert Consensus." *PLOS One* (PLOS One) 11 (8).

Erhardt, JG, JE Estes, CM Pfeiffer, HK Biesalski, and NE Craft. 2004. "Combined Measurement of Ferritin, Soluble Transferrin Receptor, Retinol Binding Protein, and C-Reactive Protein by an Inexpensive, Sensitive, and Simple Sandwich Enzyme-Linked Immunosorbent Assay Technique." *Journal of Nutrition* (134): 3127-3132.

Erhardt, JG, NE Craft, F Heinrich, and HK Biesalski. 2002. "Rapid and Simple Measurement of Retinol in Human Dried Whole Bood Spots." *Journal of Nutrition* (132): 318-321.

Garrett, DA, Sangha JK, Kothari MT, and Boyle D. 2011. "Field-friendly techniques for assessment of biomarkers of nutrition for development." *The American Journal for Clinical Nutrition* 94 (2): 685S-690S.

OIE. 2013. *Validation Guidelines.* Accessed April 8, 2019. http://www.oie.int/fileadmin/Home/eng/Health_standards/aahm/current/chapitre_validation_diagnostics_assays.pdf.

PATH. 2017. "Micronutrient Surveillance Diagnostic: Product Development Road Map Document; Target product profile for adding Folate, Vitamin B-12, and Vitamin D to an existing, commercially available micronutrient assessment tool."

1. These include the Multiple Micronutrient Assessment Tool (MMAT) (Brindle 2014)
2. Format developed to be consistent with PATH’s *Micronutrient Surveillance Diagnostic: Product Development Road Map Document; Target product profile for adding Folate, Vitamin B-12, and Vitamin D to an existing, commercially available micronutrient assessment tool.* (2017).
3. Minimal should be considered as a potential go/no-go decision point.
4. Optimistic should reflect what is needed to achieve broader, deeper, quicker global health impact; not only additional or different features than minimum.
5. Taken from BMGF presentation *Biomarkers for Micronutrient Assessment: Enough to Map Deficiency Around the World?* (6 Sep 2018).
