## Supplementary File 2 for "Target Product Profiles for a Micronutrient Assessment Tool and Associated Blood Collection Device for Use in Population-Based Surveys: An Expert Consensus"

**Supplementary File 2 (S2). Round 1 Blood Collection Device Target Product Profile (TPP) draft reviewed by the expert panel, spring 2019**

### I. Background

This blood collection device target product profile (TPP) is intended to serve as a complement to the high level “conceptual” micronutrient assessment TPP being developed to build field consensus around the best practices for use in population health surveys. As such, please reference the draft TPP for the micronutrient (MN) assessment.

### II. Target Product Profile overview**^[1]^**

|  | **Characteristic** | **Minimum (“must”)^[2]^** | **Optimal (“want”)^[3]^** | **Annotation** |
| --- | --- | --- | --- | --- |
| **1** | **Intended use** | | | |
| **1.1** | Intended use | Population surveillance of micronutrient deficiency to inform nutrition programs | Same as minimum | (PATH 2017) |
| **1.2** | Target populations | Infants/children: 6-59 months  Women of reproductive age (WRA): 15-45 years | All ages | (PATH 2017) |
| **1.3** | Target countries / Geographic coverage | Low-income or middle-income country (LMIC) | Global | (PATH 2017) |
| **1.4** | Location of use (infrastructure) | Household / field use | Same as minimum |  |
| **1.5** | Device operator: Sample collection | Trained phlebotomist | Trained household survey worker |  |
| **2** | **Device characteristics** | | | |
| 2.1 | Size and weight | Handheld | Same as minimum | (PATH 2018) |
| 2.2 | Power and water requirements | None | Same as minimum |  |
| 2.3 | Work flow requirement: throughput |  |  |  |
| 2.4 | Operating temperature | 15-35°C, 35-85% humidity | 15-40°C, 20-90% humidity | (PATH 2018) |
| 2.5 | Waste management (hazardous materials/chemicals) |  |  |  |
| 2.6 | Sterilization requirements | No additional steps required | Same as minimum |  |
| 2.7 | Consumables management |  |  |  |
| 2.8 | Additional devices or technology, such as software, required | None required | Same as minimum |  |
| 2.9 | Quality control |  |  |  |
| **3** | **Sample handling** | | | |
| 3.1 | Sample type(s) and volumes | Capillary blood  0-2 years: Recommendation needed  2-6 years: 50-100 µL (~2 drops)  >6 years: 100 µL (~4 drops) | Dried blood spot / matrix  10-40 µL (~1 drop) |  |
| 3.2 | Sample collection and transport requirements | Refrigeration required (0° C) | Ship without cold chain; should tolerate stress during transport | (PATH 2017) |
| 3.3 | Sample prep requirements | Centrifugation and aliquot required | None required |  |
| 3.4 | Operator parameters, required training |  |  |  |
| 3.5 | Sample processing requirements | In-field processing required | In-field processing not required |  |
| 3.6 | Types of specimens/assays run in the same facility |  |  |  |
| **4** | **Commercialization** | | | |
| 4.1 | Desired end-user price |  |  |  |
| 4.2 | Channels to market |  |  |  |
| 4.3 | Supply, services, and support |  |  |  |

PATH. 2018. "Assessing hemoglobinometers for maternal care: Target product profile and landscape analysis."

PATH. 2017. "Micronutrient Surveillance Diagnostic: Product Development Road Map Document; Target product profile for adding Folate, Vitamin B-12, and Vitamin D to an existing, commercially available micronutrient assessment tool."

1. Format developed to be consistent with PATH’s *Micronutrient Surveillance Diagnostic: Product Development Road Map Document; Target product profile for adding Folate, Vitamin B-12, and Vitamin D to an existing, commercially available micronutrient assessment tool.* (2017).
2. Minimal should be considered as a potential go/no-go decision point.
3. Optimistic should reflect what is needed to achieve broader, deeper, quicker global health impact; not only additional or different features than minimum.
