## Supplementary File 3 for "Target Product Profiles for a Micronutrient Assessment Tool and Associated Blood Collection Device for Use in Population-Based Surveys: An Expert Consensus"

### **Supplementary File 3 (S3). Round 2 Micronutrient Assessment Tool Target Product Profile (TPP) draft (including Round 1 results) sent to expert group for review, spring 2019**

**Rows shaded in light blue** indicate the characteristic ‘closed’ in Round 1 because it reached >75% agreement

**Blue text** indicates the Round 1 description was revised based on expert feedback

|  | **Characteristic** | **Agreement** | **Minimum – Original** | **Minimum – Revised** | **Optimal – Original** | **Optimal – Revised** | **Round 1 comments** |
| --- | --- | --- | --- | --- | --- | --- | --- |
| 1.1 | Intended use | 90% | Population surveillance of micronutrient deficiency to inform nutrition programs | None | Same as minimum | None |  |
| 1.2 | Target populations | 100%^1^ | Infants/children: 6-59 months  Women of reproductive age (WRA): 15-45 years | **Infants/children:**  **6-59 months**  **Adolescents/WRA:**  **12-49 years** | All ages | **All ages,**  **all genders** | Multiple experts comment that target populations should highlight adolescents and WRA.  Optimal standard could extend to males and screen for other health indicators, as well as extending to the elderly as a target population for Vitamin B-12, Folate, and Vitamin D. |
| 1.3 | Target countries / Geographic coverage | 100% | Low-income or middle-income country (LMIC) | None | Global | None |  |
| 1.4 | Location of use (infrastructure) | 66% | National reference laboratory | None | District-level laboratory | **Collaborative regional networks, district capacity not needed** | Multiple experts comment that large scale MN assessments are only every 5-10 years, so not worth to build capacity – either have regional collaborations or simple low-cost techniques requiring limited training.  Two experts comment that point of care device (POC)* or field clinic laboratory assessment is optimal.  ***A comment box to discuss this topic further is provided at the end of the survey.** |
| 1.5 | Assessment tool operator | 76% | Laboratorian | **Appropriately trained laboratorian** | Same as minimum | None | Two experts comment that the optimal assessment tool would be POC*  ***A comment box to discuss this topic further is provided at the end of the survey.** |
| 1.6 | Work flow requirement: throughput | 32% | 400 samples/day | **200 samples/day** | 1000 samples/day | **500 samples/day** | Multiple experts comment the need to lower the original standards to be more realistic. Also noted that high throughput not needed given low urgency and lab capacity.  Pointed out that this is dependent on lab capacity (# personnel, # assays/day)  How many samples/day does the average Demographic Heath Survey nutrition survey perform~~s~~? |
| 1.7 | Sample collection | 75% | Serum/plasma from venous blood | None | Finger/heel-prick blood-derived dried blood spot and/or serum/plasma spot | **Finger/heel-prick serum capillary sample** | Multiple experts raise that blood spots present challenges and are too limited for some assays, despite their advantages in not requiring a cold chain, recommending that small serum capillary samples are as functional and higher quality if the ultimate goal is non-invasive or less invasive assessment  Some commenters also note non-blood (saliva) or venous samples as potential alternatives. |
| 1.8 | Desired consumables storage and cold chain requirements  **(sample stability addressed in blood collection device TPP)** | 62% | 6 months at fluctuating temperatures (0-40°C);  <90% relative humidity;  no controlled temp. required | **1-2 weeks at extreme temperatures (0-30°C); Controlled temp requirement equivalent to lab requirements; No cold chain requirement** | 12 months at fluctuating temperatures (0-40°C);  No controlled temp. required;  no cold chain requirement | **Same as minimum** | Multiple experts comment that extreme temperature and time stability is not needed and 1-2 weeks is sufficient, particularly given the lab requirements for the tool; rather, more important that kit consumables are easily replaced and monitored with temperature indicator strips or kept refrigerated.  One expert noted that given long delays in survey implementation are common, longer stability can be desirable. |
| 2.1 | Format | 61% | ELISA | None | ELISA | **Low-tech bench-top analyzer with high precision** | Multiple experts comment that while ELISA is the only multiplex now, having a tool which is not AB-dependent is desirable. |
| 2.2 | Power and water requirements | 80% | Uninterrupted power and access to deionized water | Uninterrupted **daytime** power and access to deionized water | Minimal power requirements, operable from a laptop computer; plate shaker can use universal power supply | None |  |
| 2.3 | Maintenance requirements | 76% | No failures between maintenance, with maintenance scheduled annually | **No failures between maintenance with low-cost parts and regular calibration protocol;**  **equipment re-certification maintenance scheduled annually** | No failures between maintenance, with maintenance scheduled every 5 years | **No failures between maintenance, equipment re-certification maintenance scheduled every 2 years, with local maintenance technicians available** | Multiple experts comment that every 5 year maintenance is unlikely and not optimal; rather, need easy maintenance, low cost parts, regular calibration with local technicians and suggest 2 years as an acceptable optimal standard |
| 2.4 | Reagent stability | 96% | Long-term storage, requiring refrigeration | None | No refrigeration | None |  |
| 2.5 | Waste management (hazardous materials/chemicals) | 96% | Hazardous materials as needed, per WHO and country standards | None | Same as minimum | None |  |
| 2.6 | Nature of result | 57% | Qualitative;  Yes/No (deficient) indicator | **Semi-quantitative** | Quantitative | None | Multiple experts comment that changing definitions of deficient/sufficient mean that a yes/no qualitative tool is inadequate, particularly for a tool assessing multiple biomarkers; an at least-semi-quantitative tool is preferred, as quantitative results better guide intervention strategy across micronutrients. |
| 2.7 | Time to result  **(assume 150-300 samples/assay)** | 56% | 4 hours | **24 hours from sample collection** |  | **8 hours from**  **sample collection** | Multiple experts request additional clarity around this field’s parameters, pointing out the higher priority of quality and # samples/assay as additional considerations.  One expert notes that longer incubation times can increase throughput for non-individual diagnostics.  One expert notes that POC* while still in household is preferred.  ***A comment box to discuss this topic further is provided at the end of the survey.** |
| 2.8 | Ancillary supplies | 74% | Plate washer, pipettes and tips, other standard ancillary supplies necessary to performance of a lab-based immunoassay | **Determined by method: e.g. plate washer, pipettes and tips, plate reader, ancillary supplies necessary to performance of a lab-based immunoassay** | Same as minimum | None | Multiple experts comment that ancillary supplies are determined by method, asking the method for results analysis |
| 2.9 | Quality control | 90% | Standard curves fit better than 0.98; sample controls; positive/negative controls required to monitor quality of kit | **Standard curves fit better than 0.95; sample controls; positive/negative controls required to monitor quality of kit, including low and high controls** | Same as minimum | None | Multiple experts comment that a 0.98 fit is too restrictive. |
| 2.10 | Product shelf life | 85% | 12-18 months | **12 months** | 36 months (packaged with thermal indicator) | None |  |
| 2.11 | Training requirements | 78% | Recommendation needed | **Competency and equipment proficiency achievable with 2-week training** | Recommendation needed | **Competency and equipment proficiency achievable with 3-day basic training; bi-annual re-training** | Experts recommend trainings ranging from 1-2 days to 2 weeks with a frequency ranging from bi-annual (every 6 months) to annually.  Multiple experts comment on the importance of basic lab skill training and availability of tools. |
| 3.2 | Multiplexed | 78% | ≥ 1 analyte | None | ≥ 2 analytes | **≥ 5 analytes** | Multiple experts comment that the optimal standard should include more than 2 analytes |
| 3.3 | Specimen type and volume | 74% | Venipuncture  0-2 years: 70-100 µL  2-6 years: 50-100 µL (~2 drops)  >6 years: 250-500 µL (~4 drops) | **Venipuncture**  **0-2 ≤5kg: 200 mL**  **0-6 years >5kg: 500 mL**  **>6 years: 1000 mL** | Dried blood spot (DBS)/ matrix  10-40 µL (~1 drop) | **Capillary draw**  **0-6 years: 50-100 µL**  **>6 years: 250-500 µL**  **Urine sample for iodine** | Multiple experts comment that smaller volumes limit the opportunity to do necessary repeat testing while requiring the same amount of effort as a finger prick, encouraging larger volumes than DBS.  One expert recommends venipuncture to maintain greatest sample quality  One expert recommends non-blood samples (saliva) as the optimal recommendation. |
| 3.4 | **Performance range** | 94% | Limit of detection should be such that it allows clinically relevant performance as defined below | **Semi-quantitative diagnostic with cut-off value/limit** | Same as minimum | None | Multiple experts comment that clinical relevance is not the same as the provided use case (population health surveys). |
| 3.5 | Analytical specificity | 81% | 99% | None | Same as minimum | None | One expert notes that this level of specificity is not necessary. If in agreement, what level of specificity would instead be appropriate?  One expert recommends including a parameter around sensitivity as well. |
| 3.6.1 | Precision  (intra-assay variability) | 81% | 95% SD between repeats | None | Same as minimum | None |  |
| 3.6.2 | Reproducibility  (inter-assay variability) | 80% | CV < 15% | **CV < 20%** | CV < 7.5 | **CV < 15%** | Experts comment on a range of CV values from 10-20% as acceptable. |
| 3.7 | Comparative reference method | 84% | Monoplex immunoassay; HPLC; mass spectrometry | None | Same as minimum | None | Given the complexity of this topic by analyte, this field will be specifically discussed in-person at the June convening. |
| **3.8** | **Best # of repeat tests in a sample**  **(to provide confidence in the result)** |  |  | **2 tests** |  | **1 test** | This new field added as a necessary metric which will determine sample volume required and total assessment cost; added at expert recommendation. |
| 4.1 | Desired end-user price | 66% | 5 USD / analyte | **3 USD / analyte** | 1 USD / analyte | None | Multiple experts comment that 5 USD is too high for LMIC governments, also pointing out that this is dependent on the number of biomarkers in the tool. |
| 4.2 | Supply, services, and support | 94% | Ordered and supplied directly by manufacturer with global distribution and support offered via network of global distributors | None | Same as minimum | None |  |
| 4.3 | Product registration (WHO prequalification program [PQ]) | 93% | No Stringent Regulatory Authority (SRA) or WHO prequalification requirement with Research Use Only indication | None | Same as minimum | None |  |
| ^1^Although it received >75% agreement, experts suggested useful edits to the text. Thus, the field remained open for review during Round 2. | | | | | | | |

### Micronutrient Analytes/Biomarkers To Be Included in Micronutrient Assessment Tool

|  | **Minimum** | **Optimal** | **Do not include** | **Comments** |
| --- | --- | --- | --- | --- |
| Calcium | 6% | 47% | 47% | One expert notes that plasma calcium is not useful as a population assessment tool. Calcium is tightly homeostatically controlled and rarely low even during calcium deficiency.  One expert notes that this is still a research topic. |
| Iodine (Urine iodine, plasma thyroglobulin) | 70% | 30% | 0% | BOND recommended biomarker.  One expert notes this should be required for women but not children. |
| Iron (plasma ferritin) | 75% | 25% | 0% | BOND recommended biomarker.  This is noted as the best indicator, but multiple experts comment on the importance of also requiring a marker for inflammation for interpretation, requiring the addition of CRP or AGP to the list. |
| Iron (plasma soluble transferrin receptor) | 42% | 53% | 5% | Multiple experts note that this biomarker is difficult to interpret (or “terrible”), with no reference standard to quality results and increasing results with freeze thaw. |
| Iron (plasma hepcidin) | 11% | 67% | 22% | One expert notes that this has not been seen at a population level where it would add anything beyond ferritin.  One expert notes a challenge in a lack of good antibodies, but that it might be interesting to swap in instead of AGP, given it is also a good marker of inflammation. |
| Iron (plasma transferrin) | 27% | 40% | 33% | One expert notes that it is unclear how this would be interpreted. |
| Selenium (Plasma SEPP1) | 0% | 55% | 45% | One expert notes that SEPP1 (among other Selenium biomarkers) is more amenable to an ELISA-based approach than other biomarkers.  One expert notes that this is still an area of research.  One expert notes that this may only be necessary in selenium-deficient regions. |
| Vitamin A (Plasma retinol) | 48% | 48% | 5% | BOND recommended biomarker.  Multiple experts comment this is not amenable to an ELISA-based approach (requires HPLC, so not able to be done with multiple biomarkers).  One expert notes that this is a better indicator of Vitamin A status than RBP, which lacks good validation and standardization. |
| Vitamin A (Plasma RBP4) | 53% | 42% | 5% | Multiple experts comment that this would only be included as a minimum standard if retinol is not included, noting that it underestimates deficiency as there is always some in circulation (even if not bound to retinol).  One expert notes that there are still many questions about how to interpret RBP and no defined cut-off. |
| Vitamin B1 (plasma thiamine/TMP) | 21% | 57% | 21% | One expert notes that this is hard to measure.  One expert notes that this reflects recent intake and not stores nor functional outcomes, making it potentially more useful for intervention evaluation. |
| Vitamin B1 (whole blood thiamine/ TMP) | 0% | 53% | 47% | One expert notes that while having more data on B-vitamins is good, evaluation on which biomarkers to include may be more specific to vulnerable groups. |
| Vitamin B1 (RBC transkelotase) | 9% | 55% | 36% | One expert notes that a previous workshop concluded that transkelotase is probably the best biomarker because it is linked to functional outcomes, but risk of B1 deficiency is geographically and contextually specific (and this is still being defined); B1 deficiency is much lower compared to “priority” indicators routinely assessed in population-based surveys. It may not meet the criteria of either minimum not optimal, as a result. |
| Vitamin B3 (plasma niacin/nicotinamide) | 8% | 58% | 33% | One expert notes that there is no evidence for population deficiency for this micronutrient. |
| Vitamin B6 (Plasma PLP/PA) | 8% | 58% | 33% | One expert notes that there is no evidence for population deficiency for this micronutrient. |
| Vitamin B9 (RBC folate) | 27% | 63% | 11% | One expert notes that estimating population status is sufficiently done via dietary/supplement intake, but that TBD folate would to the ability to estimate folate stores. Being able to distinguish forms of folate would also be useful for distinguishing diet versus fortificant/supplement as source of folate; however, most assays are for total folate and do not allow for those distinctions.  One expert notes this should be required for women but not children. |
| Vitamin B9 (Plasma folate) | 33% | 56% | 11% | One expert notes this should be required for women but not children. |
| Vitamin B12 (Cobalamin) | 42% | 50% | 8% | One expert notes that having multiple sample types from the same sample challenges effective processing of different analytes, and there are no good antibodies for cobalamin.  One expert notes that this is sufficient to estimate B12 status at a population level, but if there were interest in characterizing degree of deficiency among individuals/groups the subsequent analytes would be useful.  One expert notes this should be required for women but not children. |
| Vitamin B12 (Plasma holotranscobalamin) | 7% | 80% | 13% | No comments given |
| Vitamin B12 (Plasma homocysteine) | 6% | 77% | 17% | One expert notes that a previous PATH TPP recorded no experts recommending HC as a biomarker, especially if there were no others included for HC. |
| Vitamin B12 (Plasma methylmalonic acid) | 15% | 64% | 22% | One expert notes that there is no good antibodies available to enable ELISA measurement of this biomarker. |
| Vitamin D (Plasma 25-OH-D) | 45% | 50% | 5% | One expert notes this is nice to have, but it is a challenge to release this from its carrier, as has been previously reported.  One expert notes that while it would be ideal to have this information (to inform prevalence of rickets in children and osteoporosis in adults), it is unclear how Vitamin D deficiency would be addressed in populations.  One expert notes that this is still an area of research. |
| Zinc (Plasma zinc) | 55% | 25% | 10% | Multiple experts comment that there is no good biomarker for zinc.  One expert notes that existing zinc kits are not effective; ICPOES are expensive, but it is possible that plasma samples could potentially be shipped to a specialty lab.  One expert notes that this difficult to collect in the field due to contamination. |
