## Supplementary File 4 for "Target Product Profiles for a Micronutrient Assessment Tool and Associated Blood Collection Device for Use in Population-Based Surveys: An Expert Consensus"

**Supplementary File 4 (S4). Round 2 Blood Collection Device Target Product Profile (TPP) draft reviewed by the expert panel (including Round 1 results), spring 2019**

**Rows shaded in light blue** indicate the characteristic ‘closed’ in Round 1 because it reached >75% agreement.

**Blue text** indicates the Round 1 description was revised based on expert feedback.

|  | **Characteristic** | **Agreement** | **Minimum – Original** | **Minimum - Revised** | **Optimal – Original** | **Optimal - Revised** | **Comments** |
| --- | --- | --- | --- | --- | --- | --- | --- |
| 1.1 | Intended use | 95% | Population surveillance of micronutrient deficiency to inform nutrition programs | **Population surveillance of micronutrient deficiency to inform nutrition programs (with possible application in intervention evaluation)** | Same as minimum | None |  |
| 1.2 | Target populations | 95%^1^ | Infants/children: 6-59 months  Women of reproductive age (WRA): 15-45 years | **Infants/children: 6-59 months**  **Adolescents/WRA: 12-49 years** | All ages | **All ages, all genders** | One expert comments that a device that works for children OR women would be an acceptable minimum standard. |
| 1.3 | Target countries / Geographic coverage | 100% | Low-income or middle-income country (LMIC) | None | Global | None |  |
| 1.4 | Location of use (infrastructure) | 100% | Household / field use | None | Same as minimum | None |  |
| 1.5 | Device operator: Sample collection | 81% | Trained phlebotomist | None | Trained household survey worker | None |  |
| 2.1 | Size and weight | 85% | Handheld | None | Same as minimum | None |  |
| 2.2 | Power and water requirements | 91% | None | None | Same as minimum | None |  |
| 2.3 | Work flow requirement: throughput  **(pre-analytical processing)** | N/a |  | **50-100 samples/day** |  | **100-200 samples/day** | Multiple experts comment on confusion on the meaning of parameter, with the need to clearly define the critical steps. Some interpreted the standard as processing mobility time (15 min to 1 hour) or time for blood collection (< 5 min/participant).  Multiple experts comment that the standard could be minimum 50/day/technician.  One expert notes that the standard could reflect phlebotomy standards in which self-collecting devices would be the optimal and fingerstick is the acceptable minimum. |
| 2.4 | Operating temperature | 72% | 15-35°C, 35-85% humidity | None | 15-40°C, 20-90% humidity | **Same as minimum** | Experts provided a broad range of recommendations, from 15-30°C for a minimum standard and up to 45°C, and one expert notes that >40°C limits the number of people around, making the high end range potentially less relevant. |
| 2.5 | Waste management (hazardous materials/chemicals) | N/a |  | **No hazardous materials or chemicals required for blood collection device; same as standard phlebotomy equipment** |  | **No hazardous materials or chemicals required for blood collection device**  **Single use and safe disposal mechanism, same as standard phlebotomy equipment** | Experts provided a broad range of suggestions for the minimum standard: No infrastructure required for use; waste management required for ancillary supplies; same disposal as standard phlebotomy equipment; single use and safe disposal mechanism.  Multiple experts comment that hazardous materials not required for blood collection device. |
| 2.6 | Consumables **stability** | N/a |  | **Stable at ambient temperature and humidity for 3 months** |  | **Stable at ambient temperature and humidity for 6 months** | Multiple experts comment that they do not understand the parameter.  Experts suggest shelf-life stability at ambient temperature or ranging from 2 weeks to 3-5 years, with one expert pointing out the consideration of gloves, swabs, collection devices, processing, and cold storage if needed. |
| 2.7 | Additional devices or technology, such as software, required | 94% | None required | None | Same as minimum | None |  |
| 2.8 | Quality control  **Sample volume validation** | N/a |  | **Clear indicator to confirm sufficient volume collected** |  | **Same as minimum** | Multiple experts comment there should be some visual indication that sufficient volume has been obtained.  Experts do not agree whether external quality control is needed or not, while some feel manufacturer built-in QC is sufficient and others recommend external QC measures.  One expert comments on the poor quality/difficulty in collecting a good DBS sample.  *Standardization added as a new field under section 3. |
| 3.1 | Sample type(s) and volumes | 78% | Capillary blood  0-2 years: Recommendation needed  2-6 years: 50-100 µL (~2 drops)  >6 years: 100 µL (~4 drops) | **Venipuncture**  **0-2 ≤5kg: 200 mL**  **0-6 years >5kg: 500 mL**  **>6 years: 1000 mL** | Dried blood spot / matrix  10-40 µL (~1 drop) | **Capillary draw**  **0-6 years: 50-100 µL**  **>6 years: 250-500 µL**  **Urine sample for iodine** | *Volumes revised to be consistent with MN Assessment Tool TPP.  Multiple experts comment that the volume should be at least 50µL if not more for all age groups, though one expert notes that without clarity around planned analytes, it is hard to determine volumes.  One expert notes that as venipuncture is safer and less painful than capillary; we should take venous blood even in small children. |
| 3.2 | Sample collection and transport requirements | 80% | Refrigeration required (0° C) | None | Ship without cold chain; should tolerate stress during transport | None |  |
| 3.3 | Sample prep requirements | 100% | Centrifugation and aliquot required | **In-field centrifugation and aliquot required** | None required | **In-field processing not required** |  |
| 3.4 | Operator parameters, required training | N/a |  | **Operator training limited, e.g. 1 day of training by competent field staff** |  | **A fully automated system where operators training isn't needed** | Expert comments range from a 2 a week minimum standard to an optimal standard ranging from 1 week to integration of a fully automated system where operators training isn't needed. |
| **3.5** | **Standardization procedures** |  |  | **Lab facility has internal standardization** |  | **Regional or global facility is accredited to conduct standardization** | New field added at expert suggestion. |
| 4.1 | Desired end-user price | N/a |  | **3 USD / sample** |  | **1 USD / sample** | Many experts agree on 1 USD; however, considerations of disposability vs reusability also noted as issues of importance.  One expert notes that the cost should be less than or equal to the price of currently available systems (e.g. lancets, disinfection material, blood collection tube). |
| 4.2 | Channels to market | N/a |  | **Mainstream regional laboratory suppliers** |  | **Mainstream regional and local laboratory suppliers** | Experts generally agree that reliable local suppliers and distributors are optimal while regional suppliers are acceptable at a minimum. |
| 4.3 | Supply, services, and support | N/a |  | **Does not require special services nor support** |  | **Same as minimum** | Multiple experts comment that supply, services, and support should not be needed if system is sufficiently robust.  One expert comments on supply chain lead times and the need for access without requiring excessive purchase quantities.  Other experts comment that manufacturers region representatives or suppliers should provide limited support and local production, with local technical capacity available from laboratories or academic institutions. |

### ^1^Although it received >75% agreement, experts suggested useful edits to the text. Thus, the field remained open for review during Round 2.
