## Supplementary File 5 for "Target Product Profiles for a Micronutrient Assessment Tool and Associated Blood Collection Device for Use in Population-Based Surveys: An Expert Consensus"

**Supplementary File 5 (S5). Round 3 Micronutrient Assessment Tool Target Product Profile (TPP) draft reviewed by the expert panel (including Round 1,2 results), summer 2019**

**Green shading:** Field closed in round 2 (R2) because it reached >75% agreement

**Light blue shading:** Field closed in round 1 (R1) because it reached >75% agreement

**Blue text** indicates the Round 2 description was revised based on expert feedback

|  | **Characteristic** | ***(Round 1) Agreement***,**%** | **(Round 2) Agreement, %** | **(Round 2) Neutral + Agreement,%** | **Minimum –**  **Round 2** | **Minimum – Revised** | **Optimal – Round 2** | **Optimal – Revised** | **Round 2 comments** |
| --- | --- | --- | --- | --- | --- | --- | --- | --- | --- |
| **SECTION 1: INTENDED USE** | | | | | | | | | |
| 1.1 | Intended use | *90%* | Closed - Not asked in Round 2 | Closed - Not asked in Round 2 | Population surveillance of micronutrient deficiency to inform nutrition programs |  | Same as minimum |  |  |
| 1.2 | Target populations | 100%^1^ | 84% | 89% | Infants/children:  6-59 months  Adolescents/women of reproductive age (WRA):  12-49 years |  | All ages,  all genders |  |  |
| 1.3 | Target countries / Geographic coverage | *100%* | Closed - Not asked in Round 2 | Closed - Not asked in Round 2 | LMIC |  | Global |  |  |
| 1.4 | Location of use (infrastructure) | *66%* | 72% | 83% | National reference laboratory |  | Collaborative regional networks, district capacity not needed | **Collaborative regional networks (centralized regional laboratory capacity)** | - One expert references the idea of "Tricorders" as in Star Trek, but recognizes point of care (POC) assessment of all biomarkers is not presently realistic. - Large-scale surveys do not need a national reference laboratory. Minimal should also be like the optimal, provided 'regional' refers to 'across several countries', e.g. West Africa, SEA, etc. - Assessment should be done every 5 years - POC/field clinic is the dream - One expert does not agree with POC, the complexities around doing this at quality are not worth the effort, it would be massively expensive to perform and what incentives are there to a company to do this? There is no high demand for such products; Optimism appreciated, but it is very ill advised to do this. - “Regional" should be clarified. Requiring that samples cross borders is not practical. - Minimum should be a regional reference laboratory (rather than a national) - Fortification programs should be monitored and evaluated much more often than every 5 years, especially initially. - Just because large scale surveys are done every 5-10 years is no reason not to build district level capacity |
| 1.5 | Assessment tool operator | *76%* | Closed - Not asked in Round 2 | Closed - Not asked in Round 2 | Appropriately trained laboratorian |  | Same as minimum |  |  |
| 1.6 | Work flow requirement: throughput | *32%* | 76% | 94% | 200 samples/day | No revision | 500 samples/day | **300 samples/day** |  |
| 1.7 | Sample collection  (Refers to method of collection rather than device used to collect sample) | *75%* | 63% | 84% | Serum/plasma from venous blood | No revision | Finger/heel-prick serum capillary sample | No revision | - Several experts agree that the current technology for dried blood spot (DBS) is not stable, it should not be ruled out given it’s potential benefits. - The change in optimal assumes a better collection device is not part of the solution. - A finger- or heel-prick is experientially less liked than venous draw. - The method for assessment should be compatible with the small sample volumes obtained by capillary blood collection, regardless of collection vessel, and believes saliva is not biologically feasible for the required biomarkers. |
| 1.8 | Desired consumables storage and cold chain requirements  (sample stability addressed in blood collection device TPP) | *62%* | 61% | 61% | 1-2 weeks at extreme temperatures (0-30°C); Controlled temp requirement equivalent to lab requirements; No cold chain requirement | **1-2 weeks at extreme temperatures (0-30°C); Controlled temp requirement equivalent to lab requirements** | Same as minimum | **1-2 months at extreme temperatures (0-30°C); Controlled temp requirement equivalent to lab requirements; No cold chain requirement** | - Many experts comment that 1-2 weeks is too short a time for extreme temperatures; longer stability should be desirable especially if assessment is to be done in country (e.g. it may not arrive before they are expired in remote area) - The "no cold chain requirement" should be moved to the optimal revised, unsure of feasibility of it as minimum |
| SECTION 2: INSTRUMENTATION | | | | | | | | | |
| 2.1 | Format | *61%* | 89% | 100% | ELISA |  | Low-tech bench-top analyzer with high precision |  |  |
| 2.2 | Power and water requirements | *80%* | Closed - Not asked in Round 2 | Closed - Not asked in Round 2 | Uninterrupted daytime power and access to deionized water |  | Minimal power requirements, operable from a laptop computer; plate shaker can use universal power supply |  |  |
| 2.3 | Maintenance requirements | *76%* | 88% | 94% | No failures between maintenance with low-cost parts and regular calibration protocol;  equipment re-certification maintenance scheduled annually |  | No failures between maintenance, equipment re-certification maintenance scheduled every 2 years, with local maintenance technicians available |  |  |
| 2.4 | Reagent stability | *96%* | Closed - Not asked in Round 2 | Closed - Not asked in Round 2 | Long-term storage, requiring refrigeration |  | No refrigeration |  |  |
| 2.5 | Waste management (hazardous materials/chemicals) | *96%* | Closed - Not asked in Round 2 | Closed - Not asked in Round 2 | Hazardous materials as needed, per WHO and country standards |  | Same as minimum |  |  |
| 2.6 | Nature of result | *57%* | 95% | 95% | Semi-quantitative |  | Quantitative |  |  |
| 2.7 | Time to result  **(from lab test initiation)**  (assume **50-100** samples/assay) | *56%* | 60% | 73% | 24 hours from sample collection | No revision | 8 hours from  sample collection | No revision | - Multiple experts note that if the assays are being done in a national reference lab, this turnaround time is unrealistic. - Minimum requirements are too stringent, as 24 hours from collection would require nearly immediate analysis after collection and efficient analytical tests; a week is recommended as minimum. - Minimum and optimal should be the same 24 hours, as there is no benefit to 8 hours since same-day results delivery to participants is not possible. - While the time to result specification is not an issue, the 150-300 samples/assay is unrealistic as the danger of sample errors increases when working with lab processes involving too many samples. |
| 2.8 | Ancillary supplies | *74%* | 94% | 94% | Determined by method: e.g. plate washer, pipettes and tips, plate reader, ancillary supplies necessary to performance of a lab-based immunoassay |  | Same as minimum |  |  |
| 2.9 | Quality control | *90%* | 94% | 100% | Standard curves fit better than 0.95; sample controls; positive/negative controls required to monitor quality of kit, including low and high controls |  | Same as minimum |  |  |
| 2.10 | Product shelf life | *85%* | Closed - Not asked in Round 2 | Closed - Not asked in Round 2 | 12 months |  | 36 months (packaged with thermal indicator) |  |  |
| 2.11 | Training requirements | *78%* | 94% | 100% | Competency and equipment proficiency achievable with 2-week training |  | Competency and equipment proficiency achievable with 3-day basic training; bi-annual re-training |  |  |
| SECTION 3: ASSESSMENT CAPABILITIES & REQUIREMENTS | | | | | | | | | |
| 3.1 | Analytes |  |  |  |  |  |  |  | **SEE NEW SECTION BELOW** |
| 3.2 | Multiplexed | *78%* | 89% | 95% | ≥ 1 analyte |  | ≥ 5 analytes |  |  |
| 3.3 | Specimen type and volume | *74%* | 84% | 89% | Venipuncture  0-2 ≤5kg: 200 µL  0-6 years >5kg: 500 µL  >6 years: 1000 µL |  | Capillary draw  0-6 years: 50-100 µL  >6 years: 250-500 µL  Urine sample for iodine |  |  |
| 3.4 | Performance range | *94%* | Closed - Not asked in Round 2 | Closed - Not asked in Round 2 | Semi-quantitative diagnostic with cut-off value/limit |  | Same as minimum |  |  |
| 3.5 | Analytical specificity | *81%* | 76% | 82% | 99% |  | Same as minimum |  | - Multiple experts comment on a potential need to set both sensitivity and specificity. without specificity, you don't know what is being measured - Several experts agree that 99% may be too restrictive as a minimal specificity, while others disagree it should be lowered. - Specificity standards may be analyte dependent. - Possibly a specificity of 95% is sufficient, but only if the sensitivity is not too low |
| 3.6.1 | Precision  (intra-assay variability) | *81%* | Closed - Not asked in Round 2 | Closed - Not asked in Round 2 | 95% SD between repeats |  | Same as minimum |  |  |
| 3.6.2 | Reproducibility  (inter-assay variability) | *80%* | 74% | 84% | CV < 20% | **CV < 15%** | CV < 15% | **CV < 10%** | *R1 Delphi originally proposed Minimum: CV<15%; Optimal CV<7.5% -- standards revised to respond to multiple expert comments in R1 (CV at LLOD use 20% and 15% as accepted norm: Tiwari G, Tiwari R. Bioanalytical method validation: An updated review. Pharm Methods. 2010;1(1):25–38.)*   - Several experts believe the revised CVs are too high and the original CVs were more appropriate - Would always be desirable to have better optimum reproducibility - One expert would have favored <15% for minimum, <10% for optimal. Problem with CV is that they ignore where the range of interest is. |
| 3.7 | Comparative reference method | *84%* | Closed - Not asked in Round 2 | Closed - Not asked in Round 2 | Monoplex immunoassay; HPLC; mass spectrometry |  | Same as minimum |  |  |
| 3.8 | Best # of repeat tests in a sample  (to provide confidence in the result) | **Field added in Round 2** | 94% | 100% | 2 tests |  | 1 test |  |  |
| SECTION 4: COMMERCIALIZATION | | | | | | | | | |
| 4.1 | Desired end-user price | *66%* | 56% | 81% | 3 USD / analyte | **5 USD / analyte** | 1 USD / analyte | No revision | - Several experts note that the minimal should be raised in order to represent a price that is both realistic and attainable, and the optimal should be more realistic so as not to give up quality. - Setting a limit on cost should not be a priority and is potentially restrictive. - Cost might vary depending on the analyte. - Cost should consider the fact that low-income and middle-income country (LMIC) governments rarely pay all the costs for a national micronutrient survey. |
| 4.2 | Supply, services, and support | *94%* | Closed - Not asked in Round 2 | Closed - Not asked in Round 2 | Ordered and supplied directly by manufacturer with global distribution and support offered via network of global distributors |  | Same as minimum |  |  |
| 4.3 | Product registration (WHO Prequalification [PQ]) | *93%* | Closed - Not asked in Round 2 | Closed - Not asked in Round 2 | No Stringent Regulatory Authority (SRA) or WHO prequalification requirement with Research Use Only indication |  | Same as minimum |  |  |

^1^Although it received >75% agreement, experts suggested useful edits to the text. Thus, the field remained open for review during Round 2.

**Section 3.1 Analytes (separated from main TPP due to different voting format)**

**Blue shading:** Field closed in Round 2 because it reached >75% agreement

**Light yellow shading:** Field achieved >50%, but less than 75%, agreement in Round 2

| 3.1.a: ≥75% agreement for micronutrient inclusion as: MINIMUM STANDARD | | | | | | |
| --- | --- | --- | --- | --- | --- | --- |
| Micronutrient | Biomarker  analyte | Rank Order Preference (Average Rank)^1^ | Minimal  (“must have”), % | Optimal  (“nice to have”), % | Do not include, % | Comments |
| Iodine | Urine iodine, Plasma thyroglobulin (TG) | N/A | 78% | 22% | 0% | - One expert notes that the minimum should be for women, and optimal for other ages - One expert says it would be ideal to have data for both women and children - One expert expresses confusion as to why the 2 biomarkers are shown together, and would prioritize TG for this tool |
| Iron |  | N/A | 100% | 0% | 0% |  |
|  | Plasma ferritin | 1 (1.18) | 94% | 6% |  | - Multiple experts comment that the minimum should be both Alpha 1-acid glycoprotein (AGP) / C-reactive protein (CRP) as well and have indicator(s) for inflammation - One expert notes that this is the best indicator - One expert notes that it should not be included (see BRINDA papers) |
|  | Plasma soluble transferrin receptor | 2 (2.43) | 24% | 65% | 12% | - One expert notes that this marker is optimal but feels it is a long way from minimum, especially with a PoC test |
|  | Plasma hepcidin | 3 (2.43) | 6% | 63% | 31% | - Multiple experts note we are far from including this in routine assays, with recent research finding Hep levels in an individual within narrow ranges, regardless of inflammatory triggers (Prentice, Science Advances 2019) and that its ability to measure inflammation differs between pregnant women and children (more data needed) - One expert notes this could be a ferritin replacement; not confounded by red blood cells (RBCs) so could use DBS for iron, iodine and vitamin A |
|  | Plasma transferrin | 4. (3.71) | 6% | 44% | 50% | No comments given |
| Vitamin A |  | N/A | 85% | 15% |  |  |
|  | Plasma retinol | 1 (1.11) | 67% | 28% | 6% | - One expert note that while not likely for multiplex, it is still a huge step forward if can measure on its own easier - One expert notes that while serum retinol is preferred over retinol binding protein (RBP), but RBP may be the only option due to analytical complications |
|  | Plasma RBP4 | 2 (1.88) | 35% | 53% | 12% | - Multiple experts say this biomarker is desired only if retinol is not available |
| ^1^ If there are multiple biomarkers available for measuring a single micronutrient, experts were asked to rank the biomarkers (analytes) from most desirable to least desirable. The final rank order presented here is based on the average rank given by experts. Biomarkers (analytes) ranked as #1 indicate it was most commonly considered the ‘best’ analyte for a given micronutrient. | | | | | | |

| 3.1.b: ≥75% Agreement for micronutrient inclusion as: OPTIMAL STANDARD | | | | | | | | | | | | |
| --- | --- | --- | --- | --- | --- | --- | --- | --- | --- | --- | --- | --- |
| Micronutrient | | Biomarker  analyte | | Rank Order Preference (Average Rank)^1^ | | Minimal  (“must have”) | | Optimal  (“nice to have”) | | Do not include | | Comments |
|  | Vitamin B1 (thiamine) |  | N/A | | 0% | | 75% | | 25% | |  | |
|  |  | Plasma thiamine/Thiamin Monophosphate (TMP) | 1  (1.24) | |  | | 69% | | 31% | | - One expert notes that given that all other markers thus far are from serum/plasma, serum/plasma has preference for logistical reasons; otherwise, this cannot be multiplexed | |
|  |  | Whole blood thiamine/TMP | 3  (2.25) | | 0% | | 54% | | 46% | | - One expert notes that as B1 is "optimal" rather than "minimal," most other assays would use plasma/serum, it would not be optimal to have whole blood assay | |
|  |  | Red blood cell (RBC) Thiamine diphosphate (ThDP) |  | | 0% | | 64% | | 36% | | No comments given | |
|  |  | RBC transkelotase | 2  (2.00) | | 8% | | 54% | | 38% | | - One expert notes that RBC-based assays should be avoided for non-"minimum " standard MN assays - One expert notes that this indicator should be included only if with other prior indication(s) of potential problems | |
|  | Vitamin D | Plasma 25-OH-D | N/A | | 17% | | 78% | | 6% | | No comments given | |
|  | Vitamin B12 |  | N/A | | 25% | | 67% | | 8% | |  | |
|  |  | Cobalamin | 1  (1.00) | | 33% | | 60% | | 7% | | - One expert agrees that serum B12 will require the same matrix and is probably sufficient for population level estimates, but that more sophisticated methods are needed for deeper digging - One expert notes that B12 deficiency is the most prevalent deficiency in the world, and that serum B12 does give picture of population status. | |
|  |  | *Plasma cobalamin* | *N/A* | | *Not included in Round 2* | | *Not included in Round 2* | | *Not included in Round 2* | |  | |
|  |  | Plasma holotranscobalamin (holoTC) | 2  (2.00) | | 0% | | 79% | | 21% | | - One expert notes that plasma holoTC in population surveys adds very little if anything to information beyond plasma B12 | |
|  |  | Plasma homocysteine (tHcy) | 3  (2.40) | | 0% | | 81% | | 19% | | - One expert notes that this isn't good on its own - One expert notes that this needs B12, B2, folate, B6 status measured as well in order to interpret high values | |
|  |  | Plasma methylmalonic acid (MMA) | 4  (3.00) | | 0% | | 67% | | 33% | | - One expert notes that this is lower priority than Hcy or MMA, especially given concerns about scale up of assay (lack of antibodies) - One expert notes that in looking at MMA EIA, we were able to find one antibody commercially available which did not recognize MMA. This is a big challenge to all EIAs, the antibodies available are really poor generally speaking. | |
|  | ^1^ If there are multiple biomarkers available for measuring a single micronutrient, experts were asked to rank the biomarkers (analytes) from most desirable to least desirable. The final rank order presented here is based on the average rank given by experts. Biomarkers (analytes) ranked as #1 indicate it was most commonly considered the ‘best’ analyte for a given micronutrient. | | | | | | | | | | | |

**3.1.c: <75% Agreement on micronutrient categorization: TO BE DISCUSSED AT CONVENING**

| Micronutrient | | Biomarker analyte | | Rank Order Preference (Average Rank)^1^ | Minimal  (“must have”) | | Optimal  (“nice to have”) | | Do not include | | Comments |
| --- | --- | --- | --- | --- | --- | --- | --- | --- | --- | --- | --- |
|  | Vitamin B3 (niacin) | | Plasma niacin/ nicotinamide | N/A | 0% | 53% | | 47% | | No comments given | |
|  | Vitamin B9 | |  | N/A | 60% | 30% | | 10% | |  | |
|  |  |  | RBC folate | 1  (1.40) | 33% | 60% | | 7% | | - One expert notes that again, for reasons of matrix and feasibility to multiplex, preference has been given to serum/plasma, even if this is a shorter-term intake - One expert notes that it is very important that either this or plasma folate be measured to monitor the effect of folic acid fortification programs, adding that many areas have incredibly high plasma folate, and folic acid addition should be reduced - One expert notes that RBC folate insufficiency is a WHO indicator for risk of neural tube defect (NTD) in women | |
|  |  |  | Plasma folate | 2  (1.60) | 36% | 50% | | 14% | | - One expert notes that it is very important that either this or RBC folate be measured to monitor the effect of folic acid fortification programs, adding that many areas have incredibly high plasma folate, and folic acid addition should be reduced | |
|  | Zinc | | Plasma zinc | N/A | 28% | 61% | | 11% | | - One expert notes that there is no good biomarker for zinc | |
|  | Calcium | | Not measured | N/A | 0% | 38% | | 63% | |  | |
|  | Vitamin B6 (Pyridoxal 5-Phosphate [PLP]) | | Plasma PLP/ Pyridoxic acid (PA) | N/A | 7% | 47% | | 47% | | - One expert notes that this should only be included in surveys if there is very specific data indicating a population/population group deficiency - One expert notes that there certainly is evidence of population deficiency of B6 | |
|  | Selenium | | Plasma Selenoprotein P (SEPP1) | N/A | 0% | 50% | | 50% | | - One expert notes that this is still an area of research, and more data is required to highlight regions where deficiency may be an issue before it is integrated into a survey MN - One expert supports inclusion for selenium-deficient regions | |

^1^ If there are multiple biomarkers available for measuring a single micronutrient, experts were asked to rank the biomarkers (analytes) from most desirable to least desirable. The final rank order presented here is based on the average rank given by experts. Biomarkers (analytes) ranked as #1 indicate it was most commonly considered the ‘best’ analyte for a given micronutrient.

**3.1.d: Removed from TPP based on Delphi Round 1 and Round 2 feedback:**

- **Choline [Plasma choline]**
- **Vitamin E [Plasma α-tocopherol]**
- **Vitamin K [PIVKA]**
- **Other B Vitamins [Plasma metabolites]**
