## Supplementary File 6 for "Target Product Profiles for a Micronutrient Assessment Tool and Associated Blood Collection Device for Use in Population-Based Surveys: An Expert Consensus"

**Supplementary File 6 (S6). Round 3 Target Product Profile (TPP) for Blood Collection Device reviewed by the expert panel (including Round 1,2 results), summer 2019**

**Green shading:** Field closed in round 2 (Round 2)

**Light blue shading:** Field closed in round 1 (R1)

**Blue text** indicates the Round 2 description was revised based on expert feedback

|  | **Characteristic** | **(Round 1) Agreement** | **(Round 2) Agreement** | **(Round 2)**  **Neutral + Agreement** | **Minimum –**  **Round 2** | **Minimum - Revised** | **Optimal – Round 2** | **Optimal - Revised** | **Round 2 comments** |
| --- | --- | --- | --- | --- | --- | --- | --- | --- | --- |
| **SECTION 1: INTENDED USE** | | | | | | | | | |
| 1.1 | Intended use | 95% | Closed - Not asked in R2 | Closed - Not asked in R2 | Population surveillance of micronutrient deficiency to inform nutrition programs (with possible application in intervention evaluation) |  | Same as minimum |  |  |
| 1.2 | Target populations | 95%^1^ | 89% | 95% | Infants/children: 6-59 months    Adolescents/WRA: 12-49 years |  | All ages, all sexes |  |  |
| 1.3 | Target countries / Geographic coverage | 100% | Closed - Not asked in R2 | Closed - Not asked in R2 | LMIC |  | Global |  |  |
| 1.4 | Location of use (infrastructure) | 100% | Closed - Not asked in R2 | Closed - Not asked in R2 | Household / field use |  | Same as minimum |  |  |
| 1.5 | Device operator: Sample collection | 81% | Closed - Not asked in R2 | Closed - Not asked in R2 | Trained phlebotomist |  | Trained household survey worker |  |  |
| **SECTION 2: DEVICE CHARACTERISTICS** | | | | | | | | | |
| 2.1 | Size and weight | 85% | Closed - Not asked in R2 | Closed - Not asked in R2 | Handheld |  | Same as minimum |  |  |
| 2.2 | Power and water requirements | 91% | Closed - Not asked in R2 | Closed - Not asked in R2 | None |  | Same as minimum |  |  |
| 2.3 | Operating temperature | 72% | 89% | 94% | 15-35°C, 35-85% humidity |  | Same as minimum |  |  |
| 2.4 | Waste management (hazardous materials/chemicals) | N/a | 89% | 100% | No hazardous materials or chemicals required for blood collection device; same as standard phlebotomy equipment |  | No hazardous materials or chemicals required for blood collection device    Single use and safe disposal mechanism, same as standard phlebotomy equipment |  |  |
| 2.5 | Consumables stability | N/a | 78% | 89% | Stable at ambient temperature and humidity for 3 months |  | Stable at ambient temperature and humidity for 6 months |  |  |
| 2.6 | Additional devices or technology, such as software, required | 94% | Closed - Not asked in R2 | Closed - Not asked in R2 | None required |  | Same as minimum |  |  |
| 2.7 | Sample volume validation | N/a | 83% | 100% | Clear indicator to confirm sufficient volume collected |  | Same as minimum |  |  |
| **SECTION 3: SAMPLE HANDING** | | | | | | | | | |
| 3.1 | Sample type(s) and volumes | 78% | 79% | 89% | Venipuncture  0-2 <5kg: 200 µL  0-6 years >5kg: 500 µL  >6 years: 1000 µL |  | Capillary draw  0-6 years: 50-100 µL  >6 years: 250-500 µL  Urine sample for iodine |  |  |
| 3.2 | Sample collection and transport requirements | 80% | Closed - Not asked in R2 | Closed - Not asked in R2 | Refrigeration required (0° C) |  | Ship without cold chain; should tolerate stress during transport |  |  |
| 3.3 | Sample prep requirements | 100% | Closed - Not asked in R2 | Closed - Not asked in R2 | In-field centrifugation and aliquot required |  | In-field processing not required |  |  |
| 3.4 | Operator parameters, required training | N/a | 72% | 78% | Operator training limited, e.g. 1 day of training by competent field staff | No revision | A fully automated system where operators training isn't needed | **Same as minimum** | - Several experts believe training should always be required and provided in some capacity for any methodology; no training is not feasible or desirable |
| **3.5** | **Standardization procedures** |  | 88% | 100% | Lab facility has internal standardization |  | Regional or global facility is accredited to conduct standardization |  |  |
| **SECTION 4: COMMERCIALIZATION** | | | | | | | | | |
| 4.1 | Desired end-user price **of device sampling** | N/a | 56% | 78% | **3 USD / sample** | No revision | **1 USD / sample** | No revision | - Multiple experts think the minimal price is too high - There is a need to consider business incentives in decision-making for the parameter, inclusive of market demand, cost of production, and product quality and comparison to market values - The minimum standard (when compared to the cost of Whatman cards at $1.40) seems high; what is realistically feasible for device manufacturers? Need to consider a market analysis. - There is confusion on if the parameter encompasses the actual cost of the collection device or cost per test. |
| 4.2 | Channels to market | N/a | 88% | 94% | Mainstream regional laboratory suppliers |  | Mainstream regional and local laboratory suppliers |  |  |
| 4.3 | Supply, services, and support | N/a | 76% | 88% | Does not require special services nor support |  | Same as minimum |  |  |

^1^Although it received >75% agreement, experts suggested useful edits to the text. Thus, the field remained open for review during Round 2.
